# Cracks in the Foundation: Disrupted Epistemic Trust as a Pathway Between Early Relational Trauma, PTSD and Disturbances in Self-Organization

**DOI:** 10.1101/2025.06.30.25330584

**Authors:** Eileen Lashani, Peter Fonagy, David Riedl, Hanna Kampling, Johannes Kruse, Astrid Lampe, Veronika Engert, P. Read Montague, Elmar Brähler, Jörg M. Fegert, Cedric Sachser, Tobias Nolte

## Abstract

Traumatic experiences profoundly impact mental and physical health, alongside an individual’s trust in their social environment. Disruptions in epistemic trust – the ability to evaluate and trust socially communicated information – are theorized to increase vulnerability to psychopathology by impairing social learning. This cross-sectional study investigated associations between trauma characteristics and epistemic stance using self-report data from a representative German sample (*N* = 2519). We examined relationships among trauma type, age at trauma, epistemic stance, PTSD and cPTSD. Neither trauma type nor timing alone predicted epistemic stance. However, their interaction revealed that early relational trauma demonstrated the strongest association with epistemic disruption. Within epistemic stance, heightened credulity mediated associations between trauma and both PTSD and cPTSD. Findings align with the hypothesis that early relational trauma disrupts epistemic trust, increasing susceptibility to psychopathology. Clarifying how trauma type and timing interact to shape epistemic stance can advance trauma-informed interventions and therapeutic approaches.

## Introduction

Traumatic events exert far-reaching effects on mental and physical health (e.g., Felitti et al., 1998; Riedl et al., 2020; McKay et al., 2021). Such events involve actual or threatened death, serious injury, sexual violence, or other forms of abuse and neglect (American Psychiatric Association, 2013). Due to their severe implications for psychosocial development, traumatic experiences significantly increase vulnerability to psychiatric disorders (McLaughlin et al., 2020). Childhood trauma, in particular, elevates the risk of depression, anxiety, disruptive behaviors, suicide attempts, substance use, cardiovascular and respiratory illnesses, cancer, and poorer overall health (Felitti et al., 1998; McLaughlin et al., 2012; Hughes et al., 2017).

A primary consequence of trauma exposure is post-traumatic stress disorder (PTSD), characterized by symptoms of re-experiencing, avoidance, negative affect, and hyperarousal (American Psychiatric Association, 2013). Closely related, complex PTSD (cPTSD) arises predominantly following severe, prolonged, or repetitive traumatization (World Health Organization, 2022). CPTSD encompasses PTSD symptoms but additionally features disturbances in self-organization (DSO), reflecting impairments in affect regulation, negative self-concept, and relational difficulties stemming from trauma-induced disruptions in personality functioning (World Health Organization, 2022). Although over 70% of individuals worldwide experience trauma, only a minority subsequently develop PTSD (Benjet et al., 2016; Liu et al., 2017). This discrepancy highlights that individual differences in resilience and vulnerability, as well as trauma characteristics, significantly modulate PTSD and cPTSD risk. The current study investigates contextual and individual factors influencing PTSD and cPTSD development, specifically examining epistemic stance as a potential mediator linking retrospectively self-reported trauma exposure to symptom severity.

Research addressing trauma characteristics suggests that the developmental timing of traumatic experiences plays a critical moderating role (Dunn et al., 2017; Teicher & Parigger, 2015). Individuals exposed to earlier traumas, particularly in early childhood, are not only significantly more likely to develop PTSD, but also to experience subsequent traumatization (Brewin et al., 2000; Dunn et al., 2017; Kessler et al., 2017; McCutcheon et al., 2010). However, evidence on the timing effect remains inconsistent, with some studies finding no significant age-related differences (Guina et al., 2018; Mueller-Pfeiffer et al., 2013), and others reporting stronger PTSD associations with later trauma onset (Copeland et al., 2007; Green et al., 1991; Schoedl et al., 2010). The biopsychosocial mechanisms underlying these heterogeneous findings are poorly understood (Dunn et al., 2017).

Additionally, studies frequently examine childhood adversity broadly, without adequately differentiating the age-dependent effects of specific trauma types (Dunn et al., 2017; Teicher & Parigger, 2015). Regarding cPTSD and associated DSO symptoms, the literature more consistently supports childhood trauma as pivotal (Cloitre et al., 2009; Karatzias et al., 2019). Nevertheless, cPTSD may also develop following adult trauma, particularly of interpersonal nature (Karatzias et al., 2019; Palic et al., 2016).

Considering that age at traumatization and trauma type may not be independent, Guina and colleagues (2018) found that the first traumatic experience is most commonly of interpersonal nature. Literature consistently indicates that interpersonal violence is particularly likely to lead to PTSD (Frans et al., 2005; Kessler et al., 2017; McLaughlin et al., 2013) and cPTSD (Karatzias et al., 2019; Palic et al., 2016). Beyond severe but less frequent interpersonal traumas such as physical assault or sexual abuse, large-scale studies have identified the unexpected death of a loved person as a prominent cause of PTSD in general populations (Breslau et al., 1998; Kessler et al., 2017). These findings suggest that not only assaultive traumas, but also relational ruptures significantly increase vulnerability to PTSD and DSO, especially when occurring early in life.

Research into mechanisms linking early adversity to psychopathology emphasizes early childhood as a critical developmental period characterized by rapid brain maturation and epigenetic programming of neuroendocrine, immune, and neurotransmitter systems, highly responsive to environmental influences (Cecil et al., 2020; Chrousos, 2009; Soares et al., 2021). Consequently, trauma occurring within this sensitive psychophysiological developmental window can have particularly disruptive effects. This period is also vital for establishing social cognition, notably attachment security, mentalizing capacity, and social learning, particularly epistemic trust (Bowlby, 1969/1982; Fonagy & Allison, 2014; Fonagy et al., 2015; Luyten et al., 2020a; Luyten et al., 2020b).

Epistemic trust, increasingly explored in recent research, describes an individual’s capacity to appraise socially communicated information as authentic, generalizable, and personally relevant (Campbell et al., 2021). Campbell and colleagues (2021) delineate three overlapping facets of epistemic stance towards socially conveyed information: trust, mistrust, and credulity. Epistemic trust represents an adaptive stance, enabling selective and appropriate engagement with social learning opportunities. Epistemic mistrust denotes generalized skepticism or rejection of socially conveyed information, viewing interpersonal communication as untrustworthy or malevolent. Epistemic credulity reflects impaired discrimination and a compromised self-position, rendering individuals susceptible to misinformation and exploitation.

The epistemic stance emerges within early attachment relationships, closely linked theoretically (Fonagy & Allison, 2014; Fonagy et al., 2015) and empirically (Campbell et al., 2021; Liotti et al., 2023; Nolte et al., under review) to attachment security and mentalizing abilities. Although disruptions in attachment and mentalizing consistently mediate the negative impact of childhood maltreatment (e.g., Li et al., 2020; Muller et al., 2012; Huang et al., 2020; Stagaki et al., 2022), recent theoretical work highlights disrupted epistemic trust as a critical third mechanism within this developmental psychopathology triad (Fonagy et al., 2017a; Fonagy et al., 2017b; Luyten et al., 2020a; Luyten et al., 2020b; Nolte et al., 2023). Fonagy and Campbell assert that “many, if not all, types of psychopathology might be characterized by temporary or permanent disruption of epistemic trust and the social learning process it enables.” (Fonagy & Campbell, 2017, p. 5).

Assuming this perspective, epistemic trust represents a critical source of resilience protecting against the development and persistence of psychopathology by facilitating social learning and enabling individuals to benefit from interpersonal relationships (Fonagy et al., 2015; Fonagy et al., 2017b; Luyten et al., 2020b). Because epistemic stance develops early in attachment relationships, traumatic relational experiences during childhood (e.g., caregiver maltreatment or loss) can disrupt adaptive epistemic trust, prompting shifts towards heightened mistrust or credulity as adaptations to a threatening interpersonal environment (Fonagy et al., 2015; Fonagy et al., 2017b; Luyten et al., 2020b). Thus, children exposed to unreliable, neglectful, or malevolent caregiving environments may become uncertain or fearful regarding others’ intentions, adopting either a hypervigilant stance of mistrust or an indiscriminate stance of credulity (Campbell et al., 2021; Nolte et al., 2023). Empirical support from cross-sectional studies increasingly aligns with this conceptualization, demonstrating negative associations between childhood adversity and epistemic trust, alongside robust positive associations with epistemic mistrust and credulity (Campbell et al., 2021; Kampling et al., 2022; Liotti et al., 2023).

Although these epistemic stances initially protect children within threatening environments, they may become maladaptive across development. Dysfunctional epistemic stances impede the growth of effective mentalizing capacities and disrupt accurate social perception and interaction in later social contexts. Trust violations during formative years or significant life events can therefore induce lasting alterations in individuals’ evaluations of trustworthiness and their acceptance of information from social agents, including friends, partners, teachers, and therapists (Fonagy et al., 2014; Nolte et al., 2023). Consequently, maladaptive epistemic stances have profound implications for psychopathology, with mistrust and credulity linked to elevated symptoms of depression, anxiety, somatization, maladaptive personality functioning, DSO, and PTSD (Campbell et al., 2021; Kampling et al., 2022; Liotti et al., 2023; Riedl et al., 2023a; Riedl et al., 2024).

Using this developmental framework, there is preliminary evidence from cross-sectional studies that a dysfunctional epistemic stance, characterized by reduced trust and elevated mistrust and credulity, acts as a mediator between childhood adversity and psychopathology (Campbell et al., 2021; Liotti et al., 2023; Nolte et al., under review), specifically concerning PTSD and DSO symptoms (Kampling et al., 2022; Riedl et al., 2024). However, these studies are limited by their cross-sectional design and reliance on general assessments of childhood trauma, often without differentiating specific trauma types or contrasting childhood trauma effects with those resulting from later-life experiences. Such comparisons are crucial to further validate the epistemic trust framework.

Taken together, trauma impacts individuals through diverse contextual and individual factors, notably age at exposure and trauma type. Given the differential associations of these trauma characteristics with PTSD and cPTSD symptomatology, this study aims to shed light on one of the potential mechanisms underlying these differences. Drawing upon theoretical frameworks and empirical findings, we propose epistemic stance as a mediator linking specific trauma characteristics to symptom severity. In line with this conceptualization, we anticipate that early relational trauma in particular undermines epistemic trust and promotes mistrust and credulity. Understanding how age at trauma exposure interacts with trauma type to influence epistemic trust, mistrust, and credulity is essential for identifying risk factors and informing trauma-informed care and interventions.

Specifically, this study aims to: 1) replicate previous findings supporting the epistemic trust framework of developmental psychopathology, whereby early adversity associates with disrupted epistemic stance and heightened psychopathology; 2) substantiate this evidence by contrasting early relational trauma with non-relational trauma across the lifespan; and 3) examine these relationships in a large representative community sample. To achieve this, we drew from a cross-sectional representative German population sample. We hypothesized that earlier traumatization (H1) and relational trauma (H2) show stronger links with epistemic stance impairments, which in turn predict increased severity of PTSD and DSO symptoms (H3).

### Transparency and Openness

The hypotheses and methods of this study were preregistered at https://osf.io/cxvej.

The analysis scripts and raw data supporting the conclusions will be made publicly available upon publication. The materials used in this study are widely accessible.

This study involved secondary analysis of existing data rather than new data collection. Power analyses were conducted to ensure the sample size was adequate for the planned analyses. Data exclusion procedures and exploratory analyses are reported transparently in the manuscript and supplementary materials.

The original data collection complied with the Declaration of Helsinki and the ethical guidelines of the International Code of Marketing and Social Research Practice of the International Chamber of Commerce and the European Society of Opinion and Marketing Research. Ethical approval was granted by the Ethics Committee of the Medical Faculty, University of Leipzig (No. 474/20-ek). Written informed consent was obtained from all participants. For minors, consent was also obtained from a parent or legal guardian.

## Method

### Participants

The present sample was drawn from a representative population survey conducted by the demographic consulting firm USUMA (Berlin, Germany) between December 2020 and March 2021. Participants were randomly selected via the ADM Sampling System, with inclusion criteria comprising age ≥ 16 years, fluency in German, and provision of written informed consent. Data were collected through face-to-face interviews and self-administered questionnaires, resulting in a final sample of *N* = 2,519 individuals (age range: 16 – 96 years; *M* = 50.33, *SD* = 18.06). The sample was 52.2% female, 47.4% male, and 0.2% identified as another gender. Regarding education, 77.3% had not completed higher education, while 21.9% had. Monthly household income was < 1,250€ for 15.2%, 1,250 – 2,500€ for 38.7%, and > 2,500€ for 46.1%. This sample has been described in greater detail in previous publications addressing different research questions (e.g., Kampling et al., 2022; Riedl et al., 2024).

### Questionnaires

#### PTSD Symptoms

To assess significant trauma and related psychopathology, the International Trauma Questionnaire (ITQ; Cloitre et al., 2018) was administered. The ITQ comprises 18 items rated on a 5-point Likert scale (1 = “not at all”, 5 = “extremely”), evaluating PTSD (re-experiencing, avoidance, sense of current threat) and DSO symptom clusters (affective dysregulation, negative self-concept, relational difficulties). While the ITQ allows categorical scoring for diagnostic purposes, we employed dimensional scores for PTSD and DSO subscales (each comprising six items; score range: 6 to 30). Internal consistencies were strong (PTSD: α = 0.89, ω = 0.94; DSO: α = 0.87, ω = 0.92).

#### Trauma Characteristics

Trauma characteristics were derived from open-ended descriptions within the ITQ, initially categorized via a semi-automated R script (version 4.4.0; R Core Team, 2024), followed by manual review. Two trained research assistants independently rated a random subset (20%), achieving excellent interrater reliability (Cohen’s κ: 0.93 – 0.99). Trauma characteristics are detailed descriptively in Table 1.

**Table 1.** Characterization of Traumatic Events.

|  | *n* | % |  | *n* | % |
| --- | --- | --- | --- | --- | --- |
| Presence of trauma |  |  | Type of trauma |  |  |
| yes | 1281 | 50.9 | loss | 512 | 20.3 |
| no | 465 | 18.5 | separation | 91 | 3.6 |
| not specified | 773 | 30.7 | emotional violence | 23 | 0.9 |
| Age at trauma |  |  | physical violence | 25 | 1.0 |
| childhood | 106 | 4.2 | sexualized violence | 34 | 1.3 |
| (≤ 18 years) | | | war/ terror | 29 | 1.2 |
| adulthood | 987 | 39.2 | natural disaster | 17 | 0.7 |
| ($>$ 18 years) | | | accident | 115 | 4.6 |
| not specified | 188 | 7.5 | illness/ surgery | 168 | 6.7 |
| Age subgroup at trauma |  |  | other | 192 | 7.6 |
| childhood | 33 | 1.3 | not specified | 75 | 3.0 |
| (≤ 12 years) | | | Nature of trauma | | |
| adolescence | 40 | 1.6 | relational | 685 | 27.2 |
| (13 – 18 years) |  |  | non-relational | 329 | 13.1 |
| young adulthood | 294 | 11.7 | not specified | 267 | 10.6 |
| (19 – 40 years) |  |  |  |  |  |
| middle adulthood | 410 | 16.3 |  |  |  |
| (41 – 65 years) |  |  |  |  |  |
| older adulthood | 152 | 6.0 |  |  |  |
| ($>$ 65 years) | | | | | |
| not specified | 352 | 14.0 |  |  |  |

*Note.* Percentages are referenced to the full sample size of $N = 2519$ .

#### Age

Using participant’s current age and indicated recency of the traumatic experience (ranging from “less than 6 months ago” to “more than 20 years ago”), the age range during traumatization was deduced. Events occurring at or before age 18 were categorized as *childhood*; events after age 18 as *adulthood*. Additionally, trauma ages were categorized according to Erikson’s developmental stages (Erikson, 1963), as displayed in Table 1. Due to low subgroup counts in some categories and based on minimum sample size requirements from an a priori power analysis, *infancy and early childhood* (< 6 years; *n* = 8) and *middle childhood* (7 to 12 years, *n* = 10) were merged into a single *childhood* category for the main analysis.

#### Trauma Type

The trauma type was determined through manual coding of participants’ trauma descriptions, based on DSM-5 (American Psychiatric Association, 2013) trauma classifications, as indicated in Table 1. The category *other* included events not fitting these classifications (e.g., COVID-19-related experiences, pet loss, economic hardship, imprisonment). Given substantial heterogeneity, the *other* category was excluded from further analyses.

#### Nature

Trauma types were dichotomized into *relational* (loss, separation, emotional, physical, sexualized violence) and *non-relational* (war and terror, natural disaster, accident, illness, surgery). Additionally, an exploratory classification differentiated trauma types as either *intentional* (emotional, physical, sexualized violence) or *non-intentional* (all remaining categories).

### Epistemic Stance

Epistemic stance was assessed using the German 12-item version of the Epistemic Trust, Mistrust and Credulity Questionnaire (ETMCQ; Campbell et al., 2021), selected due to superior validity compared to the original 15-item scale (demonstrated in the present sample; see Nolte et al., in preparation). The ETMCQ comprises three independent subscales: *trust* (five items, e.g., “I find information easier to trust and absorb when it comes from someone who knows me well.”*), mistrust* (three items, e.g., “If you put too much faith in what people tell you, you are likely to get hurt.”), and *credulity* (four items, e.g., “When I speak to different people, I find myself easily persuaded even if it is not what I believed before.”). Items were rated on a 7-point Likert scale (1 = “strongly disagree” to 7 = “strongly agree”). Internal consistencies were good for trust (α = 0.81, ω = 0.86) and credulity (α = 0.81, ω = 0.87). Given the questionable internal consistency for mistrust (α = 0.65, ω = 0.66), interpretation of results involving this subscale should be made with caution.

### Analysis

Analyses were conducted in R (version 4.4.0; R Core Team, 2024). Initially, the main analyses included only variables of primary interest; subsequently, analyses were repeated controlling for age, gender, education, and income. Participants with missing values on relevant variables were excluded from the respective analyses.

In addition to preregistered analyses, exploratory analyses were conducted, including interaction effects of age at traumatization and trauma nature on epistemic stance, a test of the full conceptual model, and sequential moderated mediation analyses. An alternative coding scheme for trauma nature was tested in supplementary analyses related to H2.

### Analysis of Variance

MANOVAs and ANOVAs were used to test construct validity and hypotheses H1 and H2. A construct check initially validated whether trauma presence (independent variable) was associated with epistemic stance (multivariate dependent variables: trust, mistrust, credulity), using a MANOVA. Criterion validity of the trauma coding was assessed via univariate ANOVAs predicting PTSD and DSO scores, and convergent validity with ACE scores was similarly tested.

Hypothesis H1 (developmental stage at trauma exposure and epistemic stance) was tested via two between-subjects MANOVAs with epistemic stance subscales as outcomes, comparing age groups (*childhood vs. adulthood vs. no trauma*) and, subsequently, detailed age subgroups. Likewise, H2 (type and nature of trauma and epistemic stance) was tested with two separate MANOVAs using trauma type (detailed subgroups) and trauma nature (*relational* vs. *non-relational* vs. *no trauma*) as independent variables. An exploratory MANOVA combining H1 and H2 evaluated interactions between age group and trauma nature.

An a priori power analysis was conducted using G*Power (version 3.1.9.6; Faul et al., 2007) to determine the required sample size for each MANOVA with three dependent variables and categorical independent variables with up to 10 levels. The analysis assumed a small-to-medium effect size (Cohen’s *f²(V)* = 0.0625), α = .05, and power (1 – β) = .90. The results indicated that a minimum of *n* = 17 per group would be necessary to detect a significant multivariate effect. Based on this, trauma age groups with insufficient sample sizes were combined to meet power requirements.

Prior to analysis, MANOVA assumptions were assessed. Univariate (values deviating from group means by ≥ 3 *SD*s) and multivariate outliers (Mahalanobis distance at α = .001) for epistemic stance were identified and excluded (*n* = 30). Post-exclusion, visual inspection indicated no problematic assumption violations.

Significant MANOVA findings were followed by univariate ANOVAs, with significant group differences explored through Tukey-Kramer HSD pairwise comparisons and Cohen’s *d* as effect sizes.

### Mediation

Two mediation models tested the link of trauma presence (binary, derived from the ITQ) with PTSD and DSO symptoms via epistemic stance (trust, mistrust, credulity) as mediators (H3). Predictors were *z*-standardized to enhance interpretability and address potential multicollinearity. Indirect effects were statistically tested with 5000 bootstrapped resamples using the psych package for R (version 2.4.6; Revelle, 2024).

### Exploratory Moderated Mediation Models

To exploratively test the full conceptual model, two moderated mediation analyses were performed (PROCESS macro for R, Model 7; Hayes, 2022). Continuous predictors were again *z*-standardized. Given that trauma characteristics only applied to participants endorsing trauma on the ITQ, analyses were limited to these individuals. Trauma nature (*relational* vs. *non-relational*) served as the independent variable, with age group moderating the paths linking trauma nature to epistemic stance (trust, mistrust, credulity). Separate models were conducted for PTSD and DSO as outcomes, respectively.

Additionally, guided by theoretical refinements after preregistration, a sequential moderated mediation model was tested exploratively. Here, trauma characteristics predicted DSO through epistemic stance, with DSO subsequently predicting PTSD symptoms. The model was implemented using custom code in PROCESS (Hayes, 2022).

## Results

Descriptive statistics and correlations among study variables are summarized in Supplementary Table 1. Results of the construct validity checks are detailed in Supplementary Note 1 and Supplementary Tables 2 and 3. In short, reporting traumatic events on the ITQ was associated with disrupted epistemic stance, particularly increased credulity. Additionally, disclosure of traumatic events appeared biased by epistemic stance, as participants withholding disclosures showed reduced epistemic trust compared to those who disclosed. Overall, the trauma coding demonstrated good criterion validity with ITQ scores and convergent validity with ACE scores.

### H1: Age at Traumatization

Results assessing effects of traumatization age are summarized in Table 2 and illustrated in Supplementary Figure 1. The MANOVA comparing traumatization in childhood, adulthood, or no trauma revealed significant differences, driven primarily by credulity.

**Table 2.**
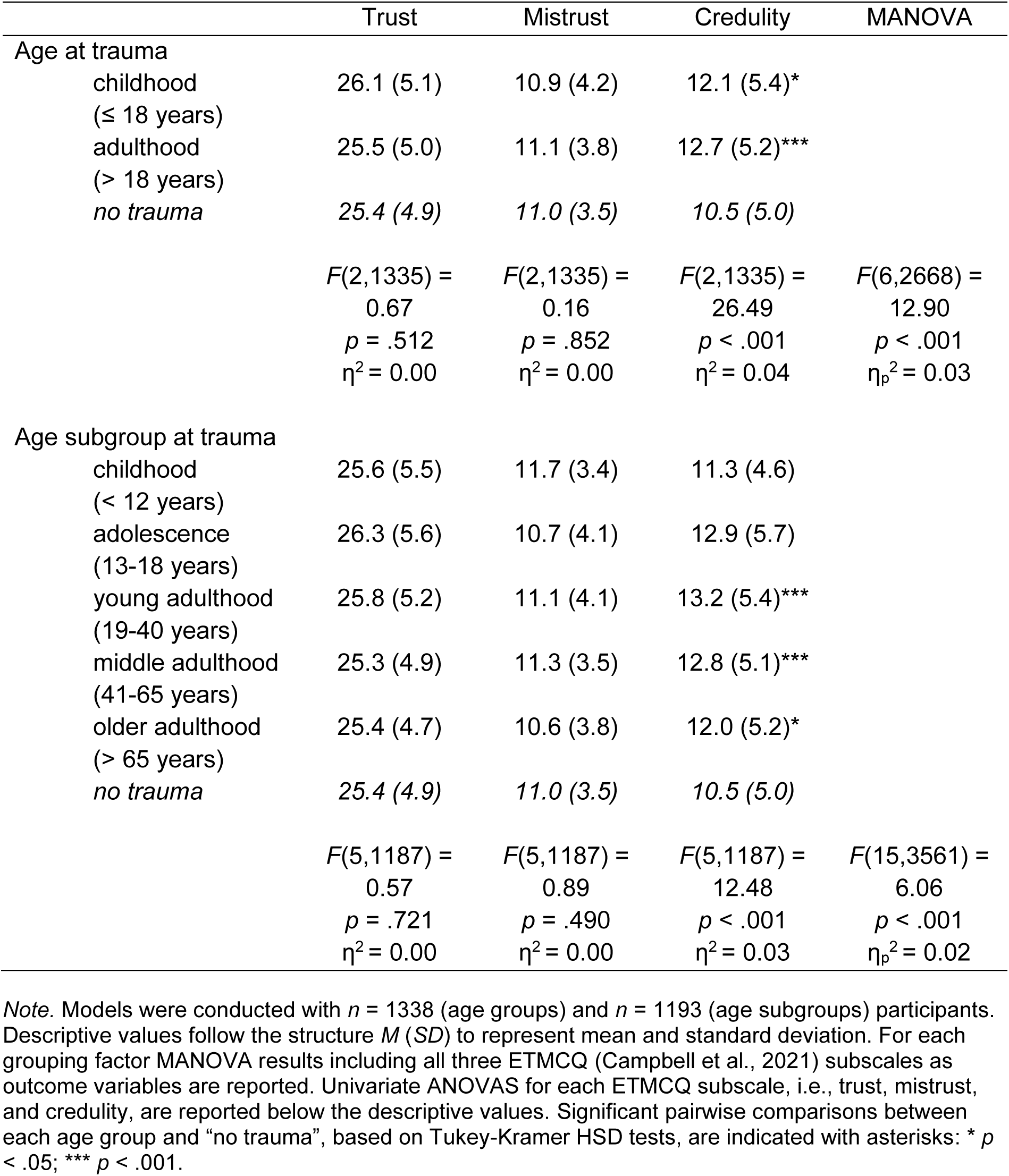
Group Differences in Epistemic Stance Between Age Groups at Traumatization.

|  | Trust | Mistrust | Credulity | MANOVA |
| --- | --- | --- | --- | --- |
| Age at trauma |  |  |  |  |
| childhood<br>($\leq 18$ years) | 26.1 (5.1) | 10.9 (4.2) | 12.1 (5.4)* | |
| adulthood<br>($> 18$ years) | 25.5 (5.0) | 11.1 (3.8) | 12.7 (5.2)*** | |
| no trauma | 25.4 (4.9) | 11.0 (3.5) | 10.5 (5.0) |  |
| | $F(2,1335) =$<br>0.67<br>$p = .512$<br>$\eta^2 = 0.00$ | $F(2,1335) =$<br>0.16<br>$p = .852$<br>$\eta^2 = 0.00$ | $F(2,1335) =$<br>26.49<br>$p < .001$<br>$\eta^2 = 0.04$ | $F(6,2668) =$<br>12.90<br>$p < .001$<br>$\eta_p^2 = 0.03$ |
| Age subgroup at trauma |  |  |  |  |
| childhood<br>($< 12$ years) | 25.6 (5.5) | 11.7 (3.4) | 11.3 (4.6) | |
| adolescence<br>(13-18 years) | 26.3 (5.6) | 10.7 (4.1) | 12.9 (5.7) |  |
| young adulthood<br>(19-40 years) | 25.8 (5.2) | 11.1 (4.1) | 13.2 (5.4)*** |  |
| middle adulthood<br>(41-65 years) | 25.3 (4.9) | 11.3 (3.5) | 12.8 (5.1)*** |  |
| older adulthood<br>($> 65$ years) | 25.4 (4.7) | 10.6 (3.8) | 12.0 (5.2)* | |
| no trauma | 25.4 (4.9) | 11.0 (3.5) | 10.5 (5.0) |  |
| | $F(5,1187) =$<br>0.57<br>$p = .721$<br>$\eta^2 = 0.00$ | $F(5,1187) =$<br>0.89<br>$p = .490$<br>$\eta^2 = 0.00$ | $F(5,1187) =$<br>12.48<br>$p < .001$<br>$\eta^2 = 0.03$ | $F(15,3561) =$<br>6.06<br>$p < .001$<br>$\eta_p^2 = 0.02$ |

*Note.* Models were conducted with $n = 1338$ (age groups) and $n = 1193$ (age subgroups) participants. Descriptive values follow the structure $M (SD)$ to represent mean and standard deviation. For each grouping factor MANOVA results including all three ETMCQ (Campbell et al., 2021) subscales as outcome variables are reported. Univariate ANOVAS for each ETMCQ subscale, i.e., trust, mistrust, and credulity, are reported below the descriptive values. Significant pairwise comparisons between each age group and “no trauma”, based on Tukey-Kramer HSD tests, are indicated with asterisks: \* $p < .05$ ; \*\*\* $p < .001$ .

Specifically, participants reporting trauma during childhood (*d* = 0.31; 95% CI [0.07, 0.55]; *p* = .033) and adulthood (*d* = 0.43; 95% CI [0.31, 0.55]; *p* < .001) both exhibited higher credulity compared to non-traumatized participants. Contrary to our hypotheses, no significant differences emerged between childhood and adulthood trauma groups in epistemic trust (*d* = −0.11; 95% CI [−0.34, 0.12]; *p* = .602), mistrust (*d* = 0.06; 95% CI [−0.17, 0.29]; *p* = .868), or credulity (*d* = 0.12; 95% CI [−0.11, 0.35]; *p* = .553).

When analyzing more refined age groups, we observed a similar pattern. Contrary to our predictions, there were no significant differences among the age subgroups in epistemic stance beyond credulity. Specifically, credulity was significantly higher among participants reporting traumatic events during young adulthood (*d* = 0.54; 95% CI [0.38, 0.70]; *p* < .001), middle adulthood (*d* = 0.47; 95% CI [0.32, 0.61]; p < .001), and older adulthood (*d* = 0.31; 95% CI [0.11, 0.51]; *p* = .032), compared to those with no reported trauma. Unexpectedly, participants who reported trauma during childhood or adolescence showed no significant differences in epistemic stance compared to those without trauma.

Given the surprising association between adulthood trauma and increased credulity, we exploratively examined whether trauma recency accounted for this finding. A multivariate linear regression indicated no significant effect of trauma recency on self-reported credulity (*b* = −0.17, *SE* = 0.11, *t*(1046) = −1.55, *p* = .122).

Including covariates (age, gender, education, income) did not alter the findings regarding age groups or subgroups at traumatization. However, we observed additional effects: older age correlated with increased mistrust and reduced credulity; male gender with lower trust and credulity; higher education with reduced credulity; and higher income with increased trust as well as decreased mistrust and credulity. Detailed covariate results are presented in Supplementary Table 4.

In summary, our hypothesis that earlier age at traumatization would be associated with greater disruption of epistemic stance was not supported. Nonetheless, credulity consistently emerged as the dimension of epistemic stance most sensitive to self-reported trauma, irrespective of age at exposure.

### H2: Type of Trauma

Results examining the effects of trauma types on epistemic stance are summarized in Table 3 and illustrated in Supplementary Figure 2. The MANOVA indicated significant differences in epistemic stance, driven primarily by credulity. Specifically, credulity was highest among participants reporting sexualized violence, significantly exceeding scores for loss (*d* = 0.62; 95% CI [0.26, 0.97]; *p* = .018) and no trauma (*d* = 0.98; 95% CI [0.62, 1.34]; *p* < .001). Other trauma types were also associated with elevated credulity compared to no trauma, including loss (*d* = 0.34; 95% CI [0.21, 0.47]; *p* < .001), separation (*d* = 0.50; 95% CI [0.26, 0.73]; *p* = .001), accidents (*d* = 0.37; 95% CI [0.16, 0.58]; *p* = .017), illness and surgery (*d* = 0.45; 95% CI [0.27, 0.64]; *p* < .001).

**Table 3.** Group Differences in Epistemic Stance Between Trauma Types.

|  | Trust | Mistrust | Credulity | MANOVA |
| --- | --- | --- | --- | --- |
| Type of trauma |  |  |  |  |
| loss | 25.3 (5.0) | 10.8 (3.7) | 12.2 (5.1)*** |  |
| separation | 25.7 (4.9) | 10.8 (3.9) | 13.0 (5.5)** |  |
| emotional violence | 24.9 (5.3) | 11.7 (3.9) | 13.3 (4.7) |  |
| physical violence | 26.2 (5.5) | 11.0 (3.8) | 13.6 (4.3) |  |
| sexualized violence | 25.9 (5.0) | 12.6 (4.1) | 15.4 (5.9)*** |  |
| war/ terror | 25.4 (6.0) | 11.0 (4.3) | 11.6 (4.4) |  |
| natural disaster | 29.2 (4.9) | 10.3 (3.4) | 12.3 (4.4) |  |
| accident | 25.3 (4.9) | 11.1 (4.1) | 12.4 (5.6)* |  |
| illness/ surgery | 25.8 (4.7) | 11.4 (3.7) | 12.7 (5.0)*** |  |
| no trauma | 25.4 (4.9) | 11.0 (3.5) | 10.5 (5.0) |  |
| | $F(9,1421) =$ | $F(9,1421) =$ | $F(9,1421) =$ | $F(27,4263) =$ |
|  | 1.42 | 1.28 | 7.26 | 4.07 |
| | $p = .172$ | $p = .242$ | $p < .001$ | $p < .001$ |
| | $\eta^2 = 0.00$ | $\eta^2 = 0.00$ | $\eta^2 = 0.04$ | $\eta_p^2 = 0.03$ |
| Nature of trauma |  |  |  |  |
| relational | 25.4 (5.0) | 10.9 (3.8) | 12.6 (5.2)*** |  |
| non-relational | 25.8 (5.0) | 11.2 (3.9) | 12.5 (5.2)*** |  |
| no trauma | 25.4 (4.9) | 11.0 (3.5) | 10.5 (5.0) |  |
| | $F(2,1428) =$ | $F(2,1428) =$ | $F(2,1428) =$ | $F(6,2854) =$ |
|  | 0.84 | 0.62 | 24.61 | 13.00 |
| | $p = .433$ | $p = .539$ | $p < .001$ | $p < .001$ |
| | $\eta^2 = 0.00$ | $\eta^2 = 0.00$ | $\eta^2 = 0.03$ | $\eta_p^2 = 0.03$ |

*Note.* Models were conducted with $n = 1431$ participants. Descriptive values follow the structure $M$ ( $SD$ ) to represent mean and standard deviation. For each grouping factor, MANOVA results including all three ETMCQ (Campbell et al., 2021) subscales as outcome variables are reported. Univariate ANOVAS for each ETMCQ subscale, i.e., trust, mistrust, and credulity, are reported below the descriptive values. Significant pairwise comparisons between each trauma type and “no trauma”, based on Tukey-Kramer HSD tests, are indicated with asterisks: \* $p < .05$ ; \*\* $p < .01$ ; \*\*\* $p < .001$ .

Examining trauma nature (*relational* vs. *non-relational*) revealed significant differences in epistemic stance, again attributable to credulity. Both relational trauma (*d* = 0.41; 95% CI [0.29, 0.53]; *p* < .001) and non-relational trauma (*d* = 0.40; 95% CI [0.25, 0.54]; *p* < .001) were associated with higher credulity compared to no trauma. However, relational and non-relational trauma did not differ significantly regarding epistemic stance.

Including covariates did not alter these patterns for trauma type or nature. Consistent with results for H1, older age correlated with increased mistrust, male gender with decreased trust and credulity, higher education with decreased credulity, and higher income with higher trust and reduced mistrust and credulity (Supplementary Table 4).

Results using the exploratory *intentional* versus *non-intentional* trauma coding scheme are detailed in Supplementary Note 2 and Supplementary Table 5. Briefly, intentional trauma was significantly associated with increased credulity compared to both non-intentional trauma and no trauma.

In summary, different trauma types were differentially related to credulity but did not show associations with trust or mistrust. Our hypothesis that relational trauma would be more detrimental to epistemic stance than non-relational trauma was not supported.

### Exploration: Interactive Effects Between Trauma Characteristics

To further elucidate how trauma characteristics jointly influence epistemic stance, we conducted an exploratory MANOVA testing the interaction between age at traumatization and trauma nature (*relational* vs. *non-relational*). This interaction significantly predicted epistemic stance (*F*[3,846] = 3.30, *p* = .020), with significant effects observed across all subscales: trust (*F*[1,848] = 5.75, *p* = .017), mistrust (*F*[1,848] = 4.76, *p* = .029), and credulity (*F*[1,848] = 4.89, *p* = .027). Main effects of age and trauma nature remained non-significant. Figure 1 illustrates these interactions, revealing consistently less disrupted epistemic stance following non-relational compared to relational trauma in childhood; this pattern did not emerge when trauma occurred later in life.

**Figure 1.**
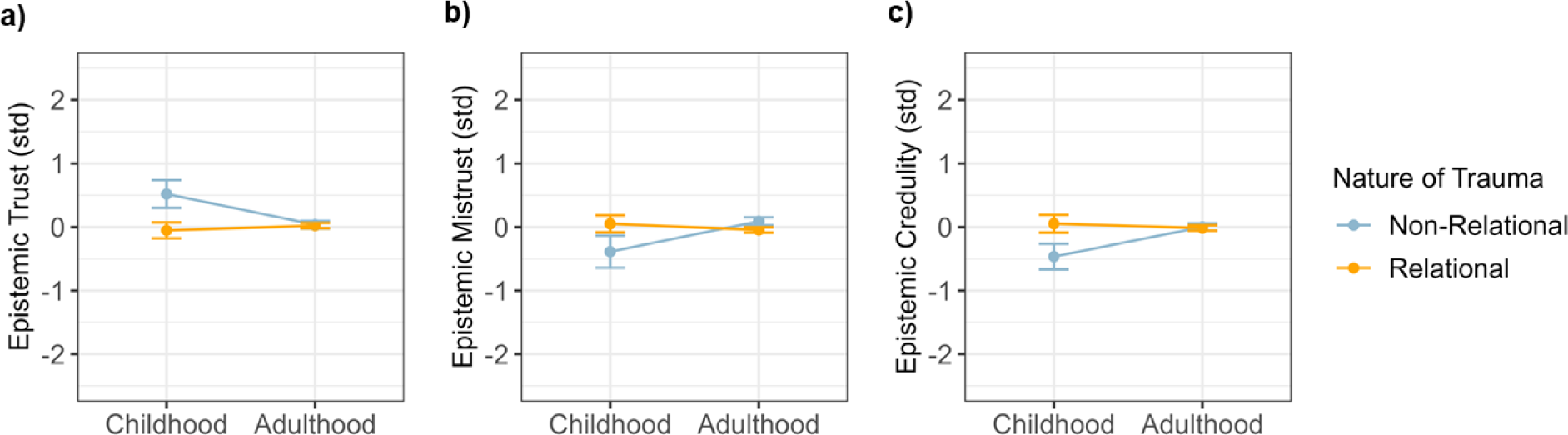
Interactions Between Nature of Trauma and Age at Occurrence. *Note.* Epistemic trust, mistrust, and credulity were assessed with the ETMCQ (Campbell et al., 2021). Original subscales range from 5 to 35, 3 to 21, and 4 to 28, respectively. Scores were z-standardized for better comparability. Childhood refers to ages ≤ 18 years, adulthood to ages > 18 years. The graphs are based on *n* = 852 participants.

Using the exploratory coding scheme (*intentional* vs. *non-intentional trauma*), no interaction effects emerged. This finding suggests that intentional trauma is broadly associated with disrupted epistemic stance across all age groups, whereas overall relational trauma effects appear age-specific (full results in Supplementary Note 2).

In summary, these exploratory findings highlight how trauma type and developmental timing interact in predicting epistemic stance disruptions, aligning with theoretical expectations.

### H3: Mediation of Trauma Effects on PTSD Symptoms Across Epistemic Stance

To test our hypothesis that epistemic stance mediates the relationship between trauma and PTSD symptoms, two mediation analyses were performed. For PTSD symptoms (*R²* = .12; *F*[4,1530] = 52.03, *p* < .001), only credulity emerged as a partial mediator between reported traumatic experience and symptom severity (β = 0.04; 95% CI [0.02, 0.05]); epistemic trust (β = 0.00; 95% CI [0.00, 0.00]) and mistrust (β = 0.00; 95% CI [0.00, 0.00]) showed no significant mediating effects (presented in Figure 2a).

**Figure 2.**
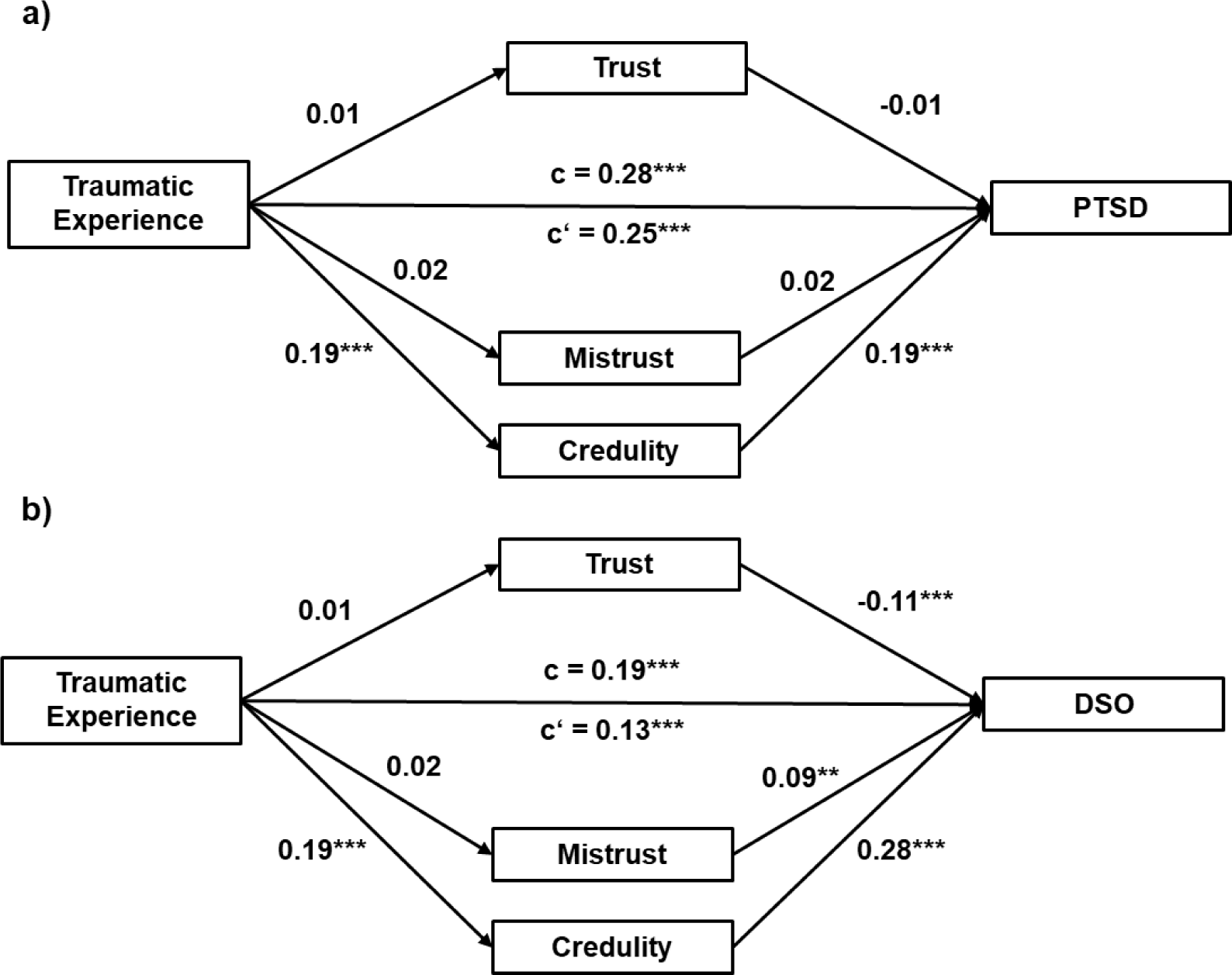
Mediation of Traumatic Experiences on PTSD and DSO Across Epistemic Stance. *Note.* The International Trauma Questionnaire (Cloitre et al., 2018) was used to assess symptoms of a) PTSD and b) disturbances in self-organization (DSO). Epistemic trust, mistrust, and credulity were assessed with the ETMCQ (Campbell et al., 2021). The graphs are based on *n* = 1535 participants. Panel a) represents the mediation model. * *p* < .05; *** *p* < .001.

For the cPTSD scale, DSO (*R²* = .17; *F*[4,1530] = 80.91, *p* < .001), we again found credulity (β = 0.05; 95% CI [0.04, 0.07]) but not trust (β = 0.00; 95% CI [−0.01, 0.01]) or mistrust (β = 0.00; 95% CI [0.00, 0.01]) to mediate symptom load linked to trauma exposure (presented in Figure 2b). Supplementary Figure 3 provides parallel graphs for additional models using ACE scores as independent variables, as preregistered and previously detailed by Kampling et al. (2022). These supplementary analyses confirmed the robustness of the mediation findings, excluding potential measurement biases from reliance on a single instrument.

In summary, credulity consistently emerged as a significant mediator linking traumatic experiences to PTSD and cPTSD symptomatology. Although mediation effects were modest, they persisted after controlling for covariates.

### Exploration: Full Conceptual Model

Testing the full conceptual model, we examined how relational trauma predicts epistemic stance depending on age at trauma, subsequently relating to PTSD and DSO symptoms. Conditional effects on mediators and conditional indirect effects on outcomes are summarized in Table 4, with full model parameters detailed in Supplementary Table 6. Figure 3 illustrates the moderated mediation model for PTSD symptoms.

**Figure 3.**
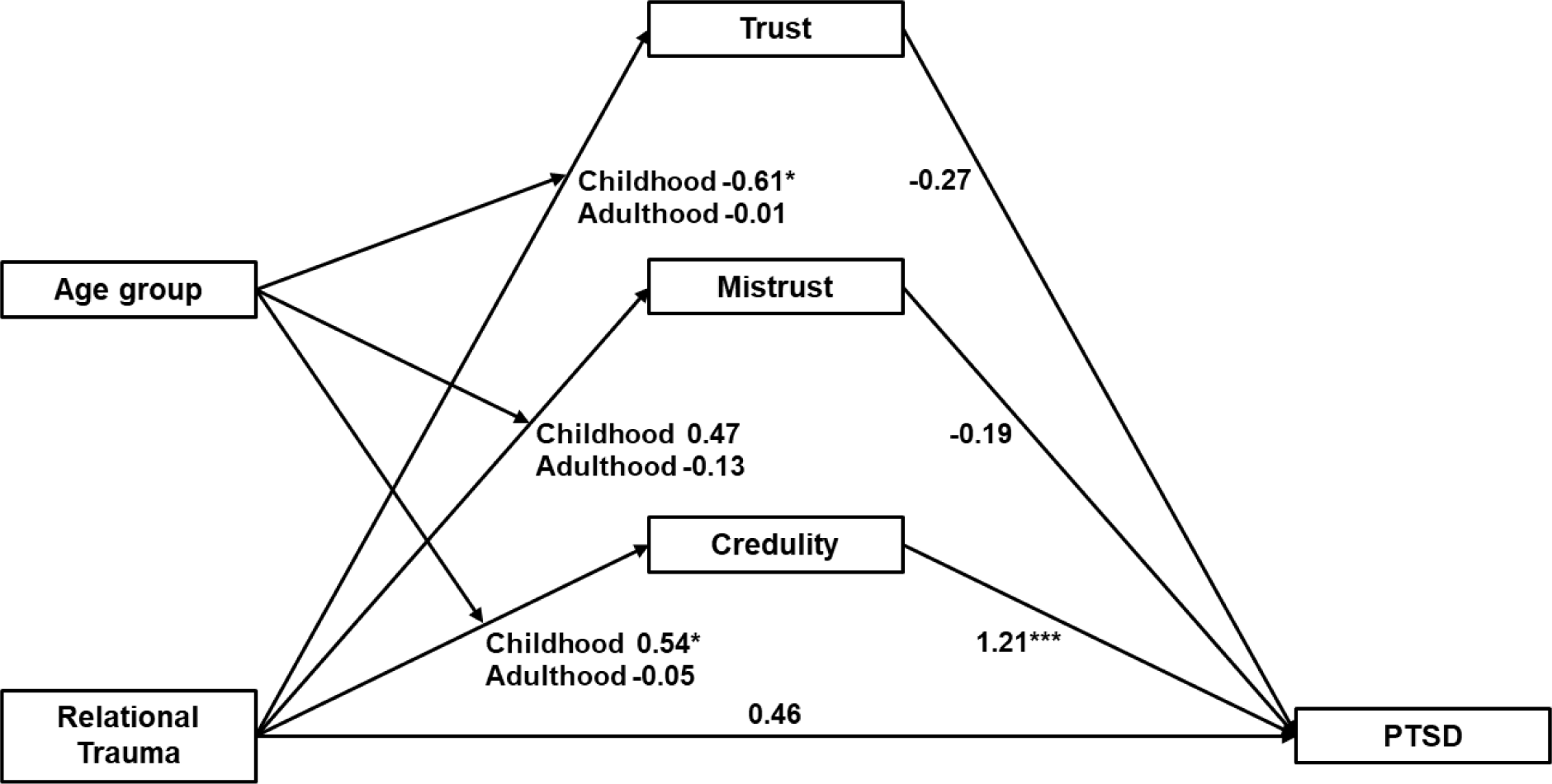
Moderated Mediation of Relational Trauma on PTSD symptoms. *Note.* The International Trauma Questionnaire (Cloitre et al., 2018) was used to assess the presence or absence of relational trauma, as well as symptoms of PTSD. Childhood refers to ages ≤ 18 years, Adulthood to ages > 18 years. Epistemic trust, mistrust, and credulity were assessed with the ETMCQ (Campbell et al., 2021). The model was conducted with *n* = 838 participants. Coefficients are standardized. * *p* < .05; *** *p* < .001.

**Table 4.**
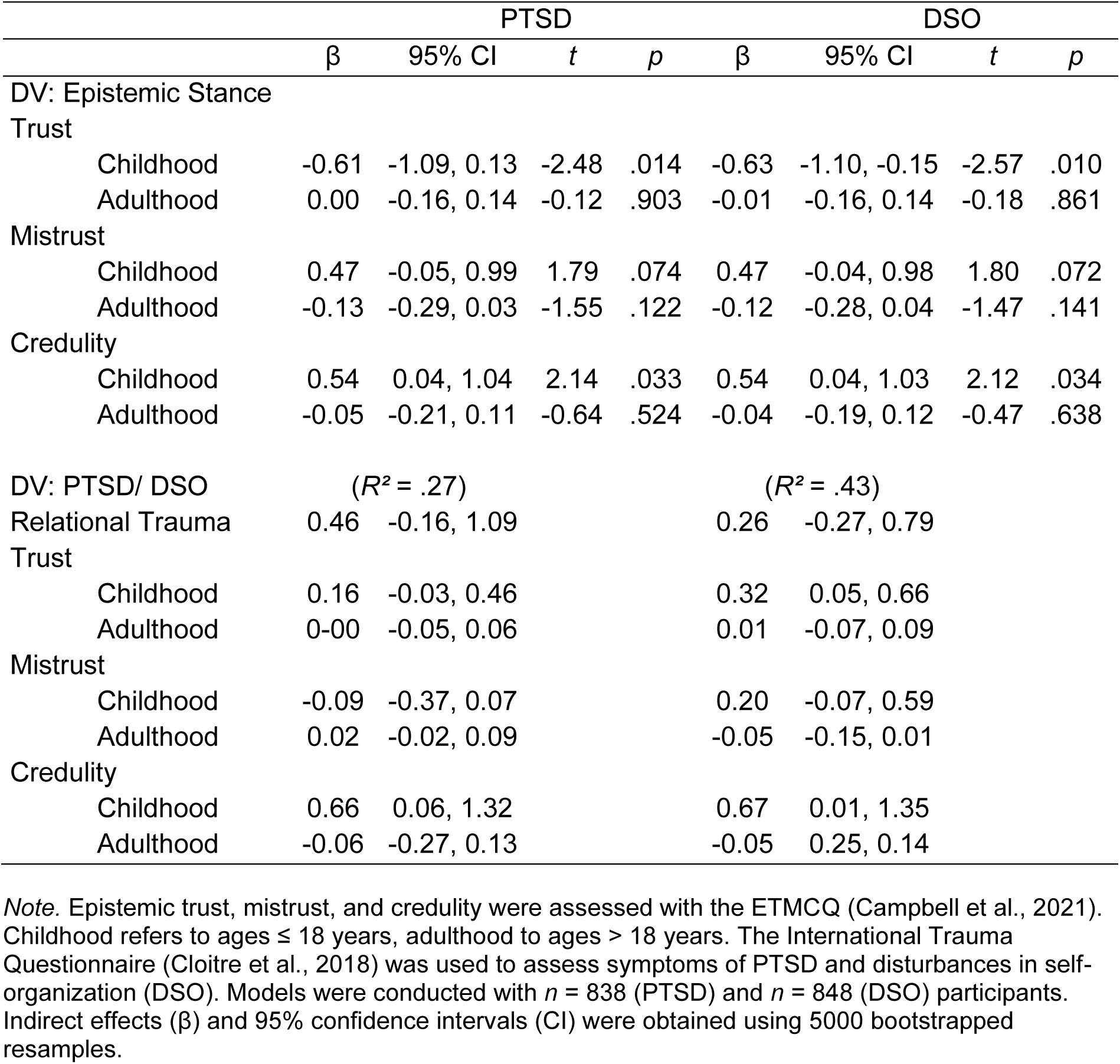
Conditional Effects of Relational Trauma on Epistemic Stance and PTSD Symptoms by Age at Traumatization.

### Effects on PTSD

Conditional effect analyses indicated that relational trauma occurring during childhood was associated with decreased epistemic trust and increased credulity. This effect was not significant for mistrust, nor were there significant effects on epistemic stance from relational trauma experienced during adulthood.

Although the direct effect of relational compared to non-relational trauma on PTSD symptoms was not significant, a significant indirect effect emerged through credulity when relational trauma occurred in childhood. The significant index of moderated mediation (IMM = −0.72, 95% CI [−1.44, −0.08]) revealed a stronger detrimental indirect effect of relational trauma on PTSD via increased credulity when trauma occurred earlier rather than later in life.

These findings suggest age at trauma moderates the indirect effect of relational trauma on PTSD symptoms through credulity.

### Effects on DSO

Similarly, relational trauma during childhood – but not adulthood – was significantly associated with decreased epistemic trust and increased credulity. Again, no significant effects were found for mistrust.

As with PTSD, relational trauma had no significant direct effect on DSO symptoms. However, significant indirect effects through epistemic trust and credulity emerged for childhood relational trauma. Both moderated mediation indices were significant (trust: IMM = −0.31, 95% CI [−0.65, −0.03]; credulity: IMM = −0.71, 95% CI [−1.42, −0.05]), indicating that relational trauma is associated with more DSO symptoms via disruptions in epistemic stance, specifically when occurring earlier in life.

Summarizing the conceptual model analyses, relational trauma was associated with disruptions in epistemic stance – particularly reduced trust and increased credulity – only when experienced during childhood. Such epistemic disruptions were, in turn, associated with elevated PTSD and DSO symptom severity.

In a final exploratory step, we tested a sequential moderated mediation model exploring whether early relational trauma and subsequent epistemic disruptions contributed to impaired self-organization, thus exacerbating PTSD symptoms. Results supported this sequential mediation pathway (fully detailed in Supplementary Note 3 and Supplementary Figure 4). In short, relational trauma in childhood predicted reduced epistemic trust and increased credulity, associated with greater DSO symptoms, which subsequently predicted heightened PTSD symptoms. Neither adulthood trauma nor the mistrust dimension demonstrated significant effects in this sequential model.

## Discussion

This study aimed to extend empirical support for the epistemic trust framework of developmental psychopathology. To this end, we built on prior research by examining the effects of trauma timing and type using a comprehensive analytic approach that accounted for a broad range of traumatic experiences across age groups. By contrasting trauma characteristics in a large representative sample of the German population, we investigated the hypotheses that early trauma (H1) and relational trauma (H2) would particularly relate to disrupted epistemic stance, and that these disruptions would mediate the relationship between trauma exposure and PTSD and cPTSD symptomatology (H3).

Our findings revealed no robust main effects for trauma timing or trauma type alone. However, a significant interaction emerged, supporting the hypothesis that early relational trauma uniquely predicts disruptions of epistemic stance. Specifically, epistemic credulity emerged as a key mediator, linking relational trauma experienced during childhood – but not adulthood – to elevated PTSD and cPTSD symptoms. Although requiring replication, these results highlight developmental specificity in trauma-related epistemic disruptions, supporting nuanced conceptualizations of how interpersonal trauma contributes to psychopathology.

The general finding that trauma is associated with disrupted epistemic stance, particularly elevated credulity, aligns with prior cross-sectional research (Campbell et al., 2021; Kampling et al., 2022; Liotti et al., 2023; Nolte et al., under review). Previous studies primarily emphasized childhood adversity; we expanded this perspective by including adult trauma. We found that both childhood and adulthood trauma were associated with elevated credulity relative to non-traumatized individuals. Contrary to our hypothesis predicting greater epistemic disruption from earlier trauma exposure, effects emerged for traumatic events during younger, middle, and older adulthood. This association could not be explained by trauma recency.

However, our trauma measure (ITQ; Cloitre et al., 2018) captured only the most salient trauma, potentially neglecting earlier experiences that profoundly shape epistemic stance. Given the cumulative nature of trauma and heightened re-traumatization risk following childhood adversity (Brewin et al., 2000; Kessler et al., 2017), early adverse experiences likely continue influencing epistemic stance beyond the explicitly reported event. Epistemic stance formation occurs early alongside other forms of social cognition (Fonagy et al., 2015; Fonagy et al., 2017a; Fonagy et al., 2017b; Luyten et al., 2020b), suggesting sustained vulnerability. Analogously, research on depression indicates that early independent stressors (e.g., abuse and neglect) increase vulnerability to subsequent dependent stressors (e.g., maladaptive relationship patterns), perpetuating maladaptive cycles of interpersonal disruption and psychopathology (Hammen, 1991; Liu & Alloy, 2010; Rudolph et al., 2000). This recursive vulnerability likely extends to epistemic disruptions, explaining their persistence even when measuring only salient traumatic events.

Consequently, inconsistent findings regarding trauma timing effects (e.g., Guina et al., 2018; Mueller-Pfeiffer et al., 2013) may stem not only from recall or measurement limitations but also from the inherent biopsychosocial complexity underlying trauma responses (Galatzer-Levy et al., 2018).

A similar pattern emerged regarding trauma type. Contrary to our second hypothesis, no consistent main effects of relational versus non-relational trauma on epistemic stance were observed. Although certain traumatic experiences, particularly sexualized violence, stood out by significantly relating to elevated credulity, the overall distinction between relational and non-relational trauma did not yield consistent differences. These findings appear to contradict prior empirical evidence linking interpersonal violence with impairments in personality functioning, mentalization, and attachment (Battle et al., 2004; Bistricky et al., 2017; Huang et al., 2017; Wagner-Skacel et al., 2022) – constructs closely related to epistemic trust (Fonagy et al., 2015; Campbell et al., 2021; Liotti et al., 2023). Using an exploratory coding approach focused on actively inflicted harm clarified that intentional trauma was associated with significantly greater credulity compared to non-intentional trauma or absence of trauma. This finding aligns with previous studies identifying assaultive interpersonal trauma as especially detrimental (Frans et al., 2005; Kessler et al., 2017; McLaughlin et al., 2013; Karatzias et al., 2019) and raises the possibility that non-intentional forms of relational trauma, such as the loss of a loved one, may have a comparatively weaker impact on epistemic trust.

Our difficulty identifying robust main effects for individual trauma characteristics mark no exception. Although factors like trauma timing and type are frequently explored predictors of PTSD, meta-analyses indicate these variables typically explain only modest variance in symptomatology (Brewin et al., 2000; Galatzer-Levy et al., 2018). Individual responses to trauma – shaped by subjective appraisal, emotional processing, and pre-existing vulnerabilities – may play a more decisive role (Ozer et al., 2003; Trickey et al., 2012; Smits et al., 2024). Such complexity could explain why our findings did not support straightforward main effects of age at trauma exposure or trauma type and instead highlight the importance of their interaction within a developmental framework.

Indeed, when combining trauma characteristics, results aligned well with our conceptual model. Specifically, relational trauma experienced in childhood, but not adulthood, was significantly associated with stronger disruptions across epistemic trust, mistrust, and credulity. Interestingly, intentional trauma showed consistently higher credulity regardless of timing, suggesting intentional interpersonal harm exerts broad detrimental effects on epistemic stance independent of age at trauma. Comparable age-insensitive effects have been documented for related outcomes, including attachment insecurity and cPTSD (Mikulincer et al., 2006; Palic et al., 2016).

Last, our hypothesis that disrupted epistemic stance mediates the association between lifetime trauma and (c)PTSD symptoms received partial support. Previous research has demonstrated mediation of childhood adversity effects on PTSD and related psychopathology via epistemic stance (Campbell et al., 2021; Kampling et al., 2022; Nolte et al., under review). Extending this, we found that epistemic credulity consistently mediated the trauma-symptom relationship across age and trauma types, with higher credulity linking trauma exposure to increased PTSD and DSO symptom severity. In contrast, neither epistemic trust nor mistrust significantly mediated these relationships. These results partially replicate earlier findings linking epistemic credulity (and mistrust) with various psychopathological outcomes, including PTSD (Campbell et al., 2021; Kampling et al., 2022; Liotti et al., 2023; Nolte et al., under review; Riedl et al., 2024).

When trauma timing and type were jointly considered, explained variance more than doubled – from 12% to 27% for PTSD, and from 17% to 43% for DSO. Supporting our hypotheses, relational trauma during childhood, but not adulthood, was significantly associated with disrupted epistemic trust and heightened credulity, which in turn predicted elevated PTSD and DSO symptoms. These findings align with developmental accounts of epistemic trust as a foundational social-cognitive mechanism shaped by early relational experiences (Fonagy et al., 2017a; Fonagy et al., 2017b; Luyten et al., 2020a; Luyten et al., 2020b; Nolte et al., 2023). Similar developmental pathways have been demonstrated for related constructs, including mentalizing, attachment, and personality functioning, linking trauma-induced disruptions to diverse psychopathological outcomes beyond PTSD and cPTSD, such as depression, anxiety, and somatic health difficulties (Li et al., 2020; Muller et al., 2012; Huang et al., 2020; Stagaki et al., 2022). To our knowledge, this study is the first to explicitly contrast childhood and adulthood trauma within the epistemic trust framework, emphasizing the unique role of early relational trauma in shaping epistemic stance and, consequently, foundational capacities for social learning and relationship formation. As indicated by exploratory sequential mediation analyses and previous research (Kampling et al., 2022), early ruptures in epistemic trust may initially impair broader personality functions (e.g., self-organization), thereby increasing vulnerability to subsequent adversity if left unaddressed through effective mentalizing interventions.

Notably, epistemic credulity emerged consistently as the dimension most sensitive to trauma across analyses, while trust and mistrust effects were less robust. It remains for future studies to explore whether epistemic credulity represents a specific psychological mechanism following trauma, whether it may be a preceding vulnerability factor, or whether this pattern reflects measurement limitations of the trust and mistrust subscales.

Integrating our findings with existing literature, it becomes clear that epistemic trust, mistrust, and credulity – constructs that have gained increasing attention over the past decade – play a key role in mental health and psychopathology. Epistemic trust is crucial for successful psychotherapy, facilitating social learning and therapeutic benefit (Fonagy et al., 2014; Nolte et al., 2023; Bröcker et al., under review). This study focused on epistemic stance in the context of trauma and (c)PTSD, conditions often marked by treatment resistance, dropout, and persistent symptoms (Bradley et al., 2005; Riedl et al., 2024; Schottenbauer et al., 2008). Identifying epistemic stance as a mechanism linking trauma to psychopathology points toward restoring epistemic trust as a promising therapeutic target – an area just beginning to be explored. Initial longitudinal evidence indicates that improvements in epistemic stance correlate with reductions in trauma-related symptoms (Riedl et al., 2023a; Riedl et al., 2023b). For instance, Lampe and colleagues (2024) recently demonstrated that increased epistemic trust and decreased credulity during psychodynamic inpatient treatment significantly predicted symptom improvement in trauma-exposed patients.

### Limitations and Directions for Future Research

A primary limitation of this study is its cross-sectional design, precluding causal interpretations of the examined pathways. Given the developmental orientation of our conceptual framework, future studies employing longitudinal designs are needed to capture temporal dynamics and developmental trajectories of epistemic trust disruptions following trauma.

While employing a large representative community sample strengthens generalizability, disrupted epistemic stance is likely even more pronounced in clinical populations. Future investigations should extend the evidence to high-risk or clinical samples. Additionally, although PTSD and DSO symptoms were analyzed dimensionally, recent preliminary research suggests caution in interpreting ITQ-derived scores without complementary clinical assessments (Shevlin et al., 2025). Moreover, as information on participants’ race and ethnicity was not available, the role of racial and ethnic diversity could not be examined, which limits the generalizability of the findings.

Another limitation concerns the suboptimal internal consistency observed for the ETMCQ mistrust scale in our sample. This aligns with prior studies reporting psychometric instability of this subscale (Asgarizadeh & Ghanbari, 2024; Campbell et al., 2021), highlighting the need for further refinement to enhance reliability.

A further challenge lies in the use of retrospective self-report for trauma assessment. Beyond the discussed limitation that the ITQ only captures the most salient traumatic event, prior evidence indicates limited overall concordance between self-reported and documented trauma exposure (Baldwin et al., 2019). Moreover, our results suggest an additional source of bias: individuals with lower epistemic trust were less likely to disclose trauma, indicating that disclosure may be shaped not only by memory but also by interpersonal openness.

### Conclusion

This study provides novel empirical support for a developmental model of epistemic trust, highlighting how early relational trauma disrupts adaptive trust in interpersonal communication, thereby contributing to trauma-related psychopathology. By systematically examining trauma timing and type, our findings demonstrate that relational context during sensitive developmental periods, rather than trauma exposure alone, critically shapes epistemic stance disruptions. Epistemic credulity, in particular, emerged as a key mechanism linking trauma to PTSD and cPTSD symptoms. These insights advance our understanding of how trauma undermines foundational social-cognitive processes and offer further indications for targeting epistemic trust in psychotherapeutic intervention efforts.

## Author Contributions

EL, TN, and PF conceptualized the project. EL, TN, PF, DR, HK, JK, AL, VE, PRM, JMF, and CS developed and executed the methodology. EL conducted the formal analysis. EB led the investigation. The original draft was written by EL, TN, and PF and critically revised by VE, DR, HK, JK, AL, PRM, EB, JMF, and CS.

## Conflicts of Interests

The authors declare that there were no conflicts of interest with respect to the authorship or the publication of this article.

## Data Availability

The analysis scripts and raw data supporting the conclusions of the present study will be made publicly available upon publication.

## Acknowledgements

We thank Alina Klettke and Amelie Sophie Schäfer for their valuable assistance with the validation of the ITQ scoring scheme. Language refinement support was provided through the use of ChatGPT (OpenAI).

## Funding

This study did not receive any specific funding from agencies in the public, commercial, or not-for-profit sectors.

## Supplemental Material

Online supplemental material is available at https://www.medrxiv.org/ along with this preprint.

## Open Science and Transparency

Hypotheses and methods of this study were preregistered at https://osf.io/cxvej. The analysis scripts and raw data supporting the conclusions will be made publicly available upon publication. All materials used in this study are widely available.

## Ethics Statement

The study was reviewed and approved by the Ethics Committee of the Medical Faculty of the University of Leipzig (474/20-ek). Written informed consent to participate in this study was provided by all participants and, if applicable, their legal guardians.

## Notes

### Competing Interest Statement

The authors have declared no competing interest.

### Author Declarations

Ethical approval was granted by the Ethics Committee of the Medical Faculty, University of Leipzig (No. 474/20-ek).

