## Supplementary Materials for "Cracks in the Foundation: Disrupted Epistemic Trust as a Pathway Between Early Relational Trauma, PTSD and Disturbances in Self-Organization"

**Supplementary Table 1**

*Descriptive Statistics and Correlations of all Included Variables*

| | *M* | *SD* | 1 | 2 | 3 | 4 | 5 | 6 | 7 | 8 | 9 | 10 |
|---|---|---|---|---|---|---|---|---|---|---|---|---|
| 1. Trauma Age | | | | | | | | | | | | |
| 2. PTSD | 8.82 | 4.19 | -.04 | | | | | | | | | |
| 3. DSO | 8.91 | 3.88 | -.09** | .52** | | | | | | | | |
| 4. Trust | 24.90 | 5.33 | -.02 | -.03 | -.18** | | | | | | | |
| 5. Mistrust | 10.97 | 3.67 | .02 | .16** | .31** | -.16** | | | | | | |
| 6. Credulity | 11.99 | 5.10 | .04 | .24** | .38** | -.14** | .57** | | | | | |
| 7. ACE | 0.90 | 1.71 | -.09** | .30** | .43** | -.11** | .18** | .26** | | | | |
| 8. Age | 50.33 | 18.06 | .45** | -.00 | .01 | -.02 | .06** | .00 | .00 | | | |
| 9. Sex (male) | | | .02 | -.10** | -.01 | -.14** | -.02 | -.10** | -.04 | -.01 | | |
| 10. Education | | | .03 | -.01 | -.05* | .02 | -.05* | -.16** | -.07** | -.11** | -.00 | |
| 11. Income | | | -.01 | -.10** | -.18** | .07** | -.13** | -.13** | -.17** | -.18** | -.08** | .16** |

*Note. M* and *SD* represent mean and standard deviation. Trauma Age refers to the age group at which trauma occurred, i.e., childhood or adulthood. The ITQ (Cloitre et al., 2018) was used to assess symptoms of PTSD and disturbances in self-organization (DSO). Epistemic trust, mistrust, and credulity were assessed with the ETMCQ (Campbell et al., 2021). Adverse childhood experiences were measured with the ACE (Schäfer et al., 2009). * $p < .05$; ** $p < .01$; *** $p < .001$.

### Supplementary Note 1

#### *Validity Check*

Epistemic stance differed between people who indicated a traumatic event, the absence of traumatic events, or chose not to make a statement. There was no difference between participants reporting the presence or absence of trauma in epistemic trust ($d = 0$; 95% CI [-0.11, 0.12]; $p = .996$) or mistrust ($d = 0.02$; 95% CI [-0.09, 0.13]; $p = .934$). Interestingly, it was those who chose not to disclose that demonstrated lower epistemic trust than participants who reported trauma ($d = -0.22$; 95% CI [-0.31, -0.12]; $p < .001$) or no trauma ($d = -0.21$; 95% CI [-0.33, -0.1]; $p = .001$). They also had lower mistrust scores than those with trauma ($d = -0.14$; 95% CI [-0.23, -0.04]; $p = .011$) but not than those without ($d = -0.12$; 95% CI [-0.24, -0.01]; $p = .118$). For credulity, people with trauma exhibited significantly higher credulity than people without trauma ($d = 0.43$; 95% CI [0.31, 0.54]; $p < .001$) and those who did not disclose ($d = 0.2$; 95% CI [0.1, 0.29]; $p < .001$). Compared to participants who reported no trauma, those who did not specify had higher credulity scores ($d = 0.25$; 95% CI [0.13, 0.36]; $p < .001$). In summary, the results of our construct check show that a traumatic event may be associated with disruptions in epistemic stance, especially credulity. Additionally, we found indications that the self-report of traumatic events may be biased by the epistemic stance.

The validity checks for our coding scheme yielded criterion validity with the ITQ. PTSD symptoms were stronger in participants with a history of trauma compared to those with no trauma ($d = 0.91$; 95% CI [0.76, 1.06]; $p < .001$) or those who did not disclose ($d = 0.89$; 95% CI [0.78, 1]; $p < .001$). There was no difference between people without trauma and those who did not specify ($d = 0.16$; 95% CI [0, 0.32]; $p = .597$). The same pattern emerged for DSO, with stronger symptoms for those with trauma than those without ($d = 0.51$; 95% CI [0.37, 0.66]; $p < .001$) or those who did not specify ($d = 0.34$; 95% CI [0.23, 0.45]; $p < .001$). People without trauma tended to display lower levels of DSO than those who did not disclose ($d = -0.22$; 95% CI [-0.38, -0.07]; $p = .061$), but this difference did not reach statistical significance.

Convergent validity with the ACE was established, showing that participants who indicated trauma on the ITQ also reported more traumatic events during childhood than participants who indicated no trauma ($d$ = 0.43; 95% CI [0.28, 0.57]; $p$ < .001) or did not disclose ($d$ = 0.39; 95% CI [0.28, 0.5]; $p$ < .001). Again, no difference was found between those who did not indicate trauma and those who did not specify ($d$ = 0.07; 95% CI [-0.09, 0.22]; $p$ = .805).

Analyses controlling for age, sex, education, and income yield the same result pattern for all models. In detail, trust was negatively associated with male gender and positively with income. Mistrust positively related to age and negatively to income. Credulity decreased with male gender, education, and income. PTSD symptoms were negatively correlated with age, male gender, and income. Both DSO and ACE scores showed negative associations with education and income.

Taken together, our construct check indicates effects of our trauma presence coding on our key epistemic stance variables and demonstrates criterion and convergent validity of our coding scheme.

**Supplementary Table 2**

*Group Differences Between Participants With and Without a Traumatic Experience*

| | Presence of trauma | | | *F* | *p* | $\eta^2$ |
|---|---|---|---|---|---|---|
| | yes | no | not disclosed | | | |
| Epistemic Stance | | | | 20.50 | < .001 | 0.03 |
| Trust | 25.4 (5.0) | 25.4 (4.9) | 24.3 (5.1) | 11.52 | < .001 | 0.01 |
| Mistrust | 11.1 (3.8) | 11.0 (3.5) | 10.6 (3.3) | 4.41 | .012 | 0.00 |
| Credulity | 12.6 (5.1) | 10.5 (5.0) | 11.7 (4.7) | 30.89 | < .001 | 0.03 |
| PTSD | 10.0 (4.4) | 6.3 (1.4) | 6.6 (1.9) | 197.50 | < .001 | 0.19 |
| DSO | 9.4 (4.1) | 7.4 (2.5) | 8.1 (3.2) | 38.14 | < .001 | 0.04 |
| ACE | 1.2 (1.9) | 0.4 (1.2) | 0.5 (1.3) | 37.34 | < .001 | 0.04 |

*Note.* Descriptive values follow the structure *M* (*SD*). MANOVA results for the epistemic stance and univariate ANOVAS for each ETMCQ (Campbell et al., 2021) subscale, i.e., trust, mistrust, and credulity are reported. Symptoms of PTSD and disturbances in self-organization (DSO) were assessed with the ITQ (Cloitre et al., 2018). Adverse childhood experiences were measured with the ACE (Schäfer et al., 2009). Models were conducted with *n* = 2235 (ETMCQ) and *n* = 1738 (other scales) participants.

**Supplementary Table 3**

*Construct, Criterion, and Convergent Validity of Trauma Coding Controlling for Covariates*

| | Trust | | | Mistrust | | | Credulity | | |
|---|---|---|---|---|---|---|---|---|---|
| | *F* | *p* | $\eta_p^2$ | *F* | *p* | $\eta_p^2$ | *F* | *p* | $\eta_p^2$ |
| Trauma | 12.71 | < .001 | 0.01 | 4.18 | .015 | 0.00 | 31.53 | < .001 | 0.03 |
| Age | 1.41 | .235 | 0.00 | 6.54 | .011 | 0.00 | 1.03 | .310 | 0.00 |
| Sex | 16.84 | < .001 | 0.02 | 0.31 | .737 | 0.00 | 9.65 | < .001 | 0.00 |
| Education | 0.11 | .745 | 0.00 | 3.55 | .060 | 0.00 | 62.70 | < .001 | 0.03 |
| Income | 4.07 | .017 | 0.00 | 9.40 | < .001 | 0.00 | 6.86 | .001 | 0.00 |
| | PTSD | | | DSO | | | ACE | | |
| | *F* | *p* | $\eta_p^2$ | *F* | *p* | $\eta_p^2$ | *F* | *p* | $\eta_p^2$ |
| Trauma | 197.69 | < .001 | 0.19 | 38.05 | < .001 | 0.04 | 37.89 | < .001 | 0.04 |
| Age | 3.84 | .050 | 0.00 | 2.15 | .143 | 0.00 | 2.97 | .085 | 0.00 |
| Sex | 5.13 | .006 | 0.00 | 0.30 | .739 | 0.00 | 0.15 | .862 | 0.00 |
| Education | 3.61 | .058 | 0.00 | 8.29 | .004 | 0.00 | 7.59 | .006 | 0.00 |
| Income | 3.05 | .048 | 0.00 | 22.97 | < .001 | 0.02 | 20.90 | < .001 | 0.02 |

*Note.* Test statistics were obtained using a MANCOVA for the epistemic stance ($F[6,4416] = 21.39$, $p < .001$, $\eta_p^2 = 0.03$) along with univariate ANCOVAs. The ETMCQ (Campbell et al., 2021) was used to measure epistemic trust, mistrust, and credulity. The ITQ (Cloitre et al., 2018) was used to assess symptoms of PTSD and disturbances in self-organization (DSO). Adverse childhood experiences were measured with the ACE (Schäfer et al., 2009). Models were conducted with *n* = 2218 (ETMCQ) and *n* = 1723 (other scales) participants.

**Supplementary Figure 1**

*Epistemic Stance by Age Group at Traumatization*

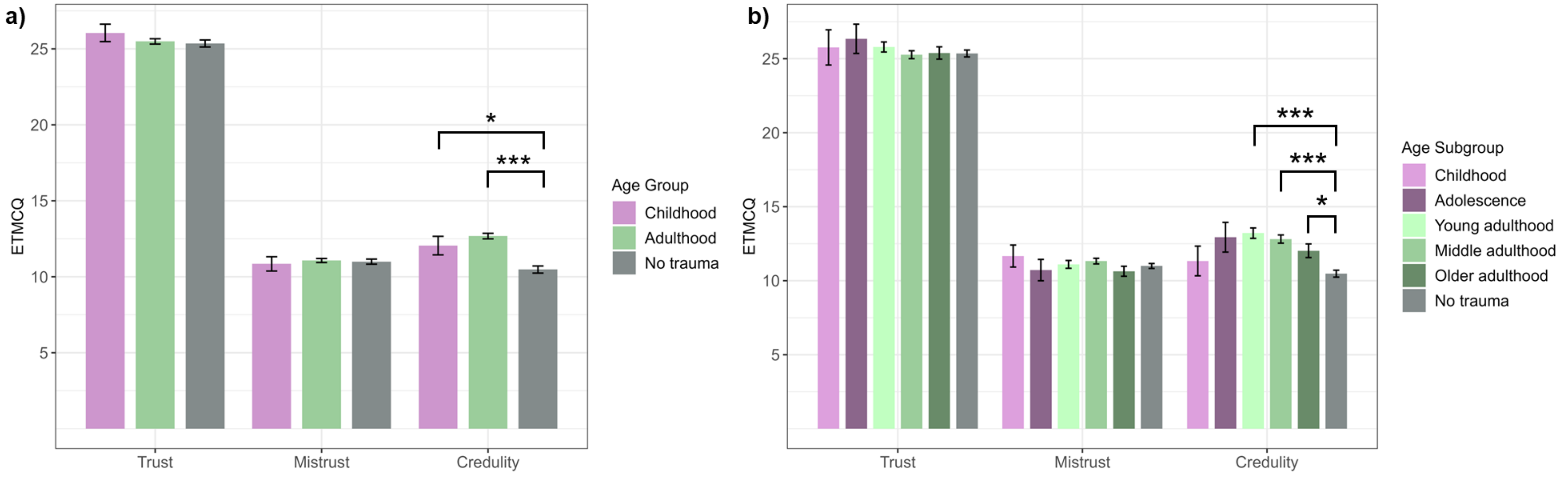

*Note.* Epistemic trust, mistrust, and credulity were assessed with the ETMCQ (Campbell et al., 2021). Subscales range from 5 to 35, 3 to 21, and 4 to 28, respectively. In panel a), childhood refers to ages ≤ 18 years, adulthood to ages > 18 years. The graph is based on $n$ = 1338 participants. In panel b), childhood refers to ages ≤ 12 years, adolescence to 13 – 18 years, young adulthood to 19 – 40 years, middle adulthood to 41 – 65 years, older adulthood to ages > 65 years. The graph is based on $n$ = 1193 participants. * $p < .05$; *** $p < .001$.

**Supplementary Table 4**

*MANCOVA Results for Group Differences in Epistemic Stance Between Age and Type Groups*

| | Trust | | | Mistrust | | | Credulity | | | MANCOVA | | |
|---|---|---|---|---|---|---|---|---|---|---|---|---|
| | *F* | *p* | $\eta_p^2$ | *F* | *p* | $\eta_p^2$ | *F* | *p* | $\eta_p^2$ | *F* | *p* | $\eta_p^2$ |
| Age at trauma | 0.64 | .528 | 0.00 | 0.17 | .845 | 0.00 | 27.00 | < .001 | 0.04 | 13.33 | < .001 | 0.03 |
| Current age | 0.48 | .489 | 0.00 | 3.06 | .081 | 0.00 | 4.74 | .030 | 0.00 | 6.51 | < .001 | 0.01 |
| Gender | 6.32 | .002 | 0.01 | 1.29 | .276 | 0.00 | 6.35 | .002 | 0.01 | 5.27 | < .001 | 0.01 |
| Education | 0.19 | .666 | 0.00 | 0.10 | .746 | 0.00 | 30.54 | < .001 | 0.02 | 14.79 | < .001 | 0.03 |
| Income | 2.26 | .105 | 0.00 | 3.28 | .038 | 0.00 | 2.37 | .093 | 0.00 | 1.72 | .112 | 0.00 |
| Age subgroup at trauma | 0.59 | .706 | 0.00 | 0.92 | .466 | 0.00 | 12.64 | < .001 | 0.05 | 6.26 | < .001 | 0.03 |
| Current age | 0.03 | .853 | 0.00 | 9.95 | .002 | 0.01 | 0.00 | .968 | 0.00 | 5.19 | .001 | 0.01 |
| Gender | 5.18 | .006 | 0.01 | 0.68 | .508 | 0.00 | 5.42 | .005 | 0.01 | 4.57 | < .001 | 0.01 |
| Education | 0.09 | .769 | 0.00 | 0.50 | .479 | 0.00 | 36.21 | < .001 | 0.03 | 16.18 | < .001 | 0.04 |
| Income | 1.85 | .158 | 0.00 | 4.20 | .015 | 0.01 | 2.83 | .060 | 0.00 | 1.99 | .064 | 0.01 |
| Trauma type | 1.40 | .182 | 0.01 | 1.32 | .224 | 0.01 | 7.54 | < .001 | 0.05 | 4.21 | < .001 | 0.03 |
| Current age | 0.24 | .627 | 0.00 | 7.13 | .008 | 0.01 | 0.45 | .501 | 0.00 | 4.96 | .002 | 0.01 |
| Gender | 8.19 | < .001 | 0.01 | 0.59 | .555 | 0.00 | 8.69 | < .001 | 0.01 | 7.34 | < .001 | 0.02 |
| Education | 0.57 | .450 | 0.00 | 0.66 | .415 | 0.00 | 34.85 | < .001 | 0.02 | 15.26 | < .001 | 0.03 |
| Income | 3.21 | .040 | 0.00 | 5.26 | .005 | 0.01 | 2.51 | .082 | 0.00 | 2.40 | .026 | 0.00 |
| Trauma nature | 0.87 | .421 | 0.00 | 0.61 | .546 | 0.00 | 25.38 | < .001 | 0.03 | 13.56 | < .001 | 0.03 |
| Current age | 0.48 | .488 | 0.00 | 4.90 | .027 | 0.00 | 1.73 | .189 | 0.00 | 5.21 | .001 | 0.01 |
| Gender | 8.73 | < .001 | 0.01 | 0.73 | .480 | 0.00 | 9.75 | < .001 | 0.01 | 7.88 | < .001 | 0.02 |
| Education | 0.49 | .483 | 0.00 | 0.62 | .433 | 0.00 | 34.55 | < .001 | 0.02 | 15.26 | < .001 | 0.03 |
| Income | 2.96 | .052 | 0.00 | 5.54 | .004 | 0.01 | 2.90 | .056 | 0.00 | 2.44 | .024 | 0.01 |

*Note.* MANCOVAs and univariate ANCOVAS for each ETMCQ (Campbell et al., 2021) subscale, i.e., trust, mistrust, and credulity, are reported. Results are reported for the grouping factors of interest (age or age subgroup at traumatization and trauma type or nature) as well as all covariates. Models were conducted with $n$ = 1330 (age group), $n$ = 1186 (age subgroup), and $n$ = 1423 (trauma type and nature) participants.

**Supplementary Figure 2**

*Epistemic Stance by Trauma Type*

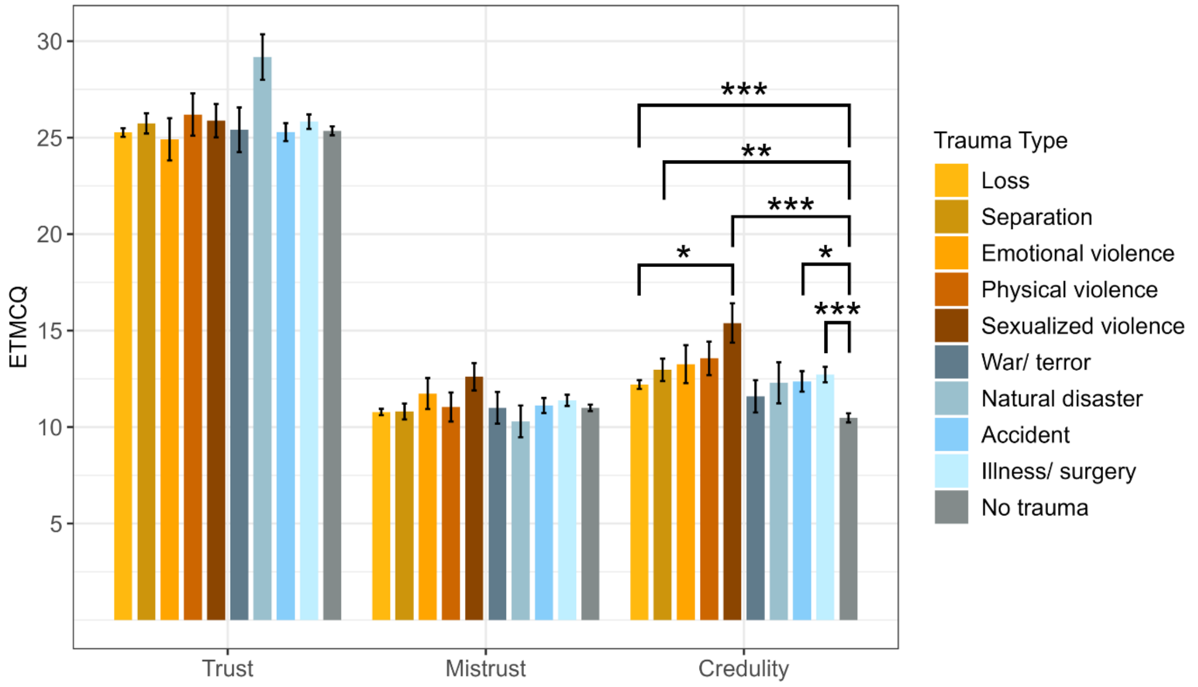

*Note.* Epistemic trust, mistrust, and credulity were assessed with the ETMCQ (Campbell et al., 2021). Subscales range from 5 to 35, 3 to 21, and 4 to 28, respectively. Trauma types marked in shades of orange belong to the category "relational trauma". Trauma types marked in shades of blue represent "non-relational trauma". The graph is based on $n$ = 1431 participants. * $p < .05$; ** $p < .01$; *** $p < .001$.

**Supplementary Note 2**

***H2: Type of Trauma Using an Alternative Coding Scheme***

Based on theoretical considerations after preregistration, a second dichotomization was explored, distinguishing trauma that was intentionally inflicted on an individual by another person (i.e., emotional, physical, and sexualized violence; $n$ = 82; 3.3%), and non-intentional trauma (i.e., loss, separation, war and terror, natural disaster, accident, illness and surgery; $n$ = 932; 37.0%).

Therefore, in H2, a third MANOVA tested intentional vs. non-intentional vs. no trauma as a grouping factor to predict epistemic stance. When grouping the trauma types by intentionally inflicted harm by another person or non-intentional traumatization, univariate differences in credulity were observed. While both intentional ($d$ = 0.75; 95% CI [0.51, 0.99]; $p < .001$) and unintentional ($d$ = 0.37; 95% CI [0.26, 0.49]; $p < .001$) traumatization were again associated with increased credulity compared to no trauma, participants that reported intentional traumatization also demonstrated significantly higher credulity than those with other traumas ($d$ = 0.36; 95% CI [0.13, 0.59]; $p$ = .005). In conclusion, these additional analyses show that it was the intentional rather than relational nature of the trauma, which was associated with increased credulity.

***Exploration: Interactive Effects Between Trauma Characteristics Using an Alternative Coding Scheme***

We also conducted an exploratory MANOVA testing the interaction between age at traumatization and the intentional vs. non-intentional nature of the trauma. This interaction did not reach significance for the multivariate model ($F$[3,846] = 1.88, p = .131), nor for any of the univariate analyses (Trust: $F$[1,848] = 3.77, p = .053; Mistrust: $F$[1,848] = 2.89, p = .089; Credulity: $F$[1,848] = 0.59, p = .444), even if trends for trust and mistrust could be noted. Instead, the main effect of intentionality remained the dominant predictor of epistemic stance ($F$[3,846] = 3.17, p = .024). This indicates that intentional trauma impacts the epistemic stance in any age group, whereas the effects of broader relational trauma are more age group specific.

**Supplementary Table 5**

*Group Differences in Epistemic Stance Between Intentional and Non-Intentional Trauma*

| | Trust | Mistrust | Credulity | MANOVA |
|---|---|---|---|---|
| Nature of trauma | | | | |
| intentional | 25.5 (5.0) | 10.9 (3.4) | 12.4 (5.2)*** | |
| non-intentional | 25.7 (5.2) | 11.9 (3.9) | 14.2 (5.1)*** | |
| *no trauma* | *25.4 (4.9)* | *11.0 (3.5)* | *10.5 (5.0)* | |
| | $F$(2,1428) = 0.22<br>$p$ = .805<br>$\eta_p^2$ = 0.00 | $F$(2,1428) = 2.43<br>$p$ = .088<br>$\eta_p^2$ = 0.00 | $F$(2,1428) = 29.65<br>$p$ < .001<br>$\eta_p^2$ = 0.04 | $F$(6,2854) = 14.10<br>$p$ < .001<br>$\eta_p^2$ = 0.03 |

| | Trust | Mistrust | Credulity | MANCOVA |
|---|---|---|---|---|
| Nature of trauma | $F$(2,1414) = 0.21<br>$p$ = .808<br>$\eta_p^2$ = 0.00 | $F$(2,1414) = 2.48<br>$p$ = .084<br>$\eta_p^2$ = 0.00 | $F$(2,1414) = 30.82<br>$p$ < .001<br>$\eta_p^2$ = 0.04 | $F$(6,2826) = 14.79<br>$p$ < .001<br>$\eta_p^2$ = 0.03 |
| Current age | $F$(1,1414) = 0.48<br>$p$ = .489<br>$\eta_p^2$ = 0.00 | $F$(1,1414) = 6.45<br>$p$ = .011<br>$\eta_p^2$ = 0.00 | $F$(1,1414) = 0.69<br>$p$ = .405<br>$\eta_p^2$ = 0.00 | $F$(3,1412) = 4.95<br>$p$ = .002<br>$\eta_p^2$ = 0.01 |

| | | | | |
|---|---|---|---|---|
| Gender | $F(2,1414) = 8.31$<br>$p < .001$<br>$\eta_p^2 = 0.01$ | $F(2,1414) = 0.51$<br>$p = .600$<br>$\eta_p^2 = 0.00$ | $F(2,1414) = 8.94$<br>$p < .001$<br>$\eta_p^2 = 0.01$ | $F(6,2826) = 7.46$<br>$p < .001$<br>$\eta_p^2 = 0.02$ |
| Education | $F(1,1414) = 0.41$<br>$p = .522$<br>$\eta_p^2 = 0.00$ | $F(1,1414) = 0.64$<br>$p = .423$<br>$\eta_p^2 = 0.00$ | $F(1,1414) = 34.23$<br>$p < .001$<br>$\eta_p^2 = 0.02$ | $F(3,1412) = 15.01$<br>$p < .001$<br>$\eta_p^2 = 0.03$ |
| Income | $F(2,1414) = 3.06$<br>$p = .047$<br>$\eta_p^2 = 0.00$ | $F(2,1414) = 5.17$<br>$p = .006$<br>$\eta_p^2 = 0.01$ | $F(2,1414) = 2.76$<br>$p = .064$<br>$\eta_p^2 = 0.00$ | $F(6,2826) = 2.35$<br>$p = .029$<br>$\eta_p^2 = 0.01$ |

*Note.* Models were conducted with $n = 1431$ (no covariates) and $n = 1423$ (including covariates) participants. Descriptive values follow the structure *M* (*SD*) to represent mean and standard deviation. For each grouping factor MANOVA results including all three ETMCQ (Campbell et al., 2021) subscales as outcome variables are reported. Univariate ANOVAS for each ETMCQ subscale, i.e., trust, mistrust, and credulity, are reported below the descriptive values. Significant pairwise comparisons between each trauma type and "no trauma", based on Tukey-Kramer HSD tests, are indicated with asterisks: *** p < .001.

**Supplementary Figure 3**

*Mediation Models of ACEs on PTSD and DSO Through Epistemic Stance*

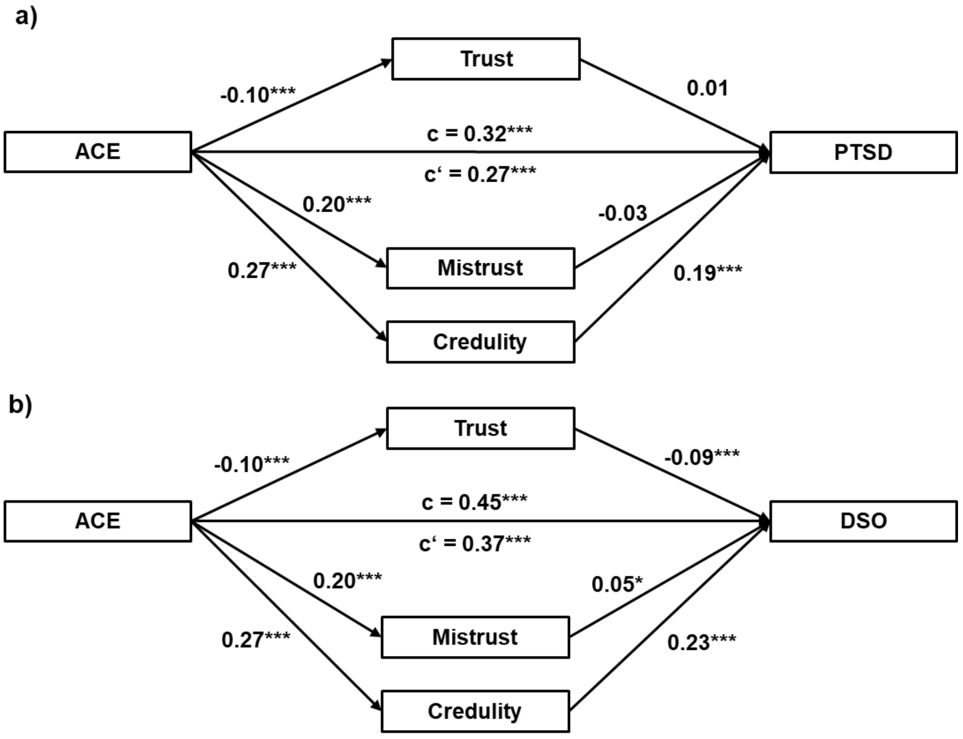

*Note.* The ITQ (Cloitre et al., 2018) was used to assess symptoms of a) PTSD and b) disturbances in self-organization (DSO). Adverse childhood experiences were measured with the ACE (Schäfer et al., 2009). Epistemic trust, mistrust, and credulity were assessed with the ETMCQ (Campbell et al., 2021). Models were conducted with n = 1535 participants. * $p < .05$; ***; $p < .001$.

**Supplementary Table 6**

*Moderated Mediation Results of Relational Trauma Through Epistemic Stance on PTSD and DSO by Age Group*

| | PTSD | | | | DSO | | | |
|---|---|---|---|---|---|---|---|---|
| | β | 95% CI | *t* | *p* | β | 95% CI | *t* | *p* |
| DV: Trust | | | | | | | | |
| Intercept | 1.14 | 0.35, 1.93 | 2.83 | .005 | 1.14 | 0.35, 1.93 | 2.84 | .005 |
| Relational Trauma | -1.20 | -2.18, -0.23 | -2.43 | .015 | -1.24 | -2.21, -0.27 | -2.51 | .012 |
| Age Group | -0.51 | -0.92, -0.10 | -2.46 | .014 | -0.52 | -0.93, -0.11 | -2.48 | .013 |
| Relational Trauma:Age Group | 0.60 | 0.09, 1.10 | 2.33 | .020 | 0.61 | 0.11, 1.11 | 2.40 | .017 |
| DV: Mistrust | | | | | | | | |
| Intercept | -0.91 | -1.76, -0.06 | -2.10 | .036 | -0.89 | -1.74, -0.05 | -2.07 | .039 |
| Relational Trauma | 1.07 | 0.02, 2.11 | 2.01 | .045 | 1.06 | 0.02, 2.09 | 2.01 | .045 |
| Age Group | 0.52 | 0.08, 0.96 | 2.31 | .021 | 0.50 | 0.06, 0.94 | 2.24 | .025 |
| Relational Trauma:Age Group | -0.60 | -1.14, -0.06 | -2.17 | .030 | -0.59 | -1.13, -0.05 | -2.16 | .031 |
| DV: Credulity | | | | | | | | |
| Intercept | -0.90 | -1.72, -0.07 | -2.13 | .033 | -0.88 | -1.70, -0.06 | -2.10 | .036 |
| Relational Trauma | 1.14 | 0.13, 2.15 | 2.21 | .028 | 1.11 | 0.11, 2.12 | 2.17 | .030 |
| Age Group | 0.53 | 0.11, 0.96 | 2.46 | .014 | 0.52 | 0.09, 0.94 | 2.39 | .017 |
| Relational Trauma:Age Group | -0.60 | -1.12, -0.07 | -2.23 | .026 | -0.57 | -1.09, -0.05 | -2.17 | .031 |
| DV: PTSD/ DSO | | | | | | | | |
| Intercept | 9.77 | 9.25, 10.28 | 37.14 | < .001 | 9.07 | 8.63, 9.51 | 40.56 | < .001 |
| Relational Trauma | 0.46 | -0.16, 1.09 | 1.45 | .148 | 0.26 | -0.27, 0.79 | 0.96 | .339 |
| Trust | -0.27 | -0.57, 0.03 | -1.75 | .080 | -0.51 | -0.76, -0.25 | -3.88 | < .001 |
| Mistrust | -0.19 | -0.54, 0.16 | -1.04 | .297 | 0.44 | 0.14, 0.73 | 2.86 | .004 |
| Credulity | 1.21 | 0.85, 1.57 | 6.67 | < .001 | 1.24 | 0.94, 1.55 | 8.04 | < .001 |

*Note.* All models were conducted with a subsample of $n$ = 838 participants. Epistemic trust, mistrust, and credulity were assessed with the ETMCQ (Campbell et al., 2021). The ITQ (Cloitre et al., 2018) was used to assess symptoms of PTSD and disturbances in self-organization (DSO). Parameters were determined using Model 7 in PROCESS (Hayes, 2022).

**Supplementary Note 3**

***Exploration: Sequential Moderated Mediation***

Based on theoretical considerations after preregistration, we tested a sequential moderated mediation. The model used the effects of trauma and its characteristics to predict DSO through epistemic stance. DSO, in turn, predicted the severity of PTSD symptoms. The model was implemented in PROCESS (Hayes, 2022) using custom code.

The direct effect of relational trauma on PTSD was not significant (β = 0.39, 95% CI [-0.16, 0.94], $t(832) = 1.40$, $p = .163$), neither was the indirect effect of relational trauma on PTSD through DSO (β = 0.15, 95% CI [-0.16, 0.44]).

When looking at conditional effects, the indirect effect of relational trauma on PTSD through Trust and DSO was significant if trauma occurred in childhood (β = 0.17, 95% CI [0.02, 0.36]), but not significant if trauma occurred during adulthood (β = 0.00, 95% CI [-0.04, 0.05]). The corresponding moderated mediation effect was also significant (IMM = -0.16, 95% CI [-0.37, -0.02]).

Similarly, the indirect effect of relational trauma on PTSD through Credulity and DSO was significant if trauma occurred in childhood (β = 0.39, 95% CI [0.02, 0.79]), but not significant if trauma occurred during adulthood (β = -0.02, 95% CI [-0.14, 0.09]). The corresponding moderated mediation effect was also significant (IMM = -0.42, 95% CI [-0.83, -0.04]).

However, the indirect effect of relational trauma on PTSD through Mistrust and DSO was not significant irrespective of whether trauma occurred during childhood (β = 0.12, 95% CI [-0.03, 0.34]) or adulthood (β = -0.03, 95% CI [-0.09, 0.01]).

**Supplementary Figure 4**

*Sequential Moderated Mediation of Relational Trauma on PTSD Through Epistemic Stance by Age Group and DSO*

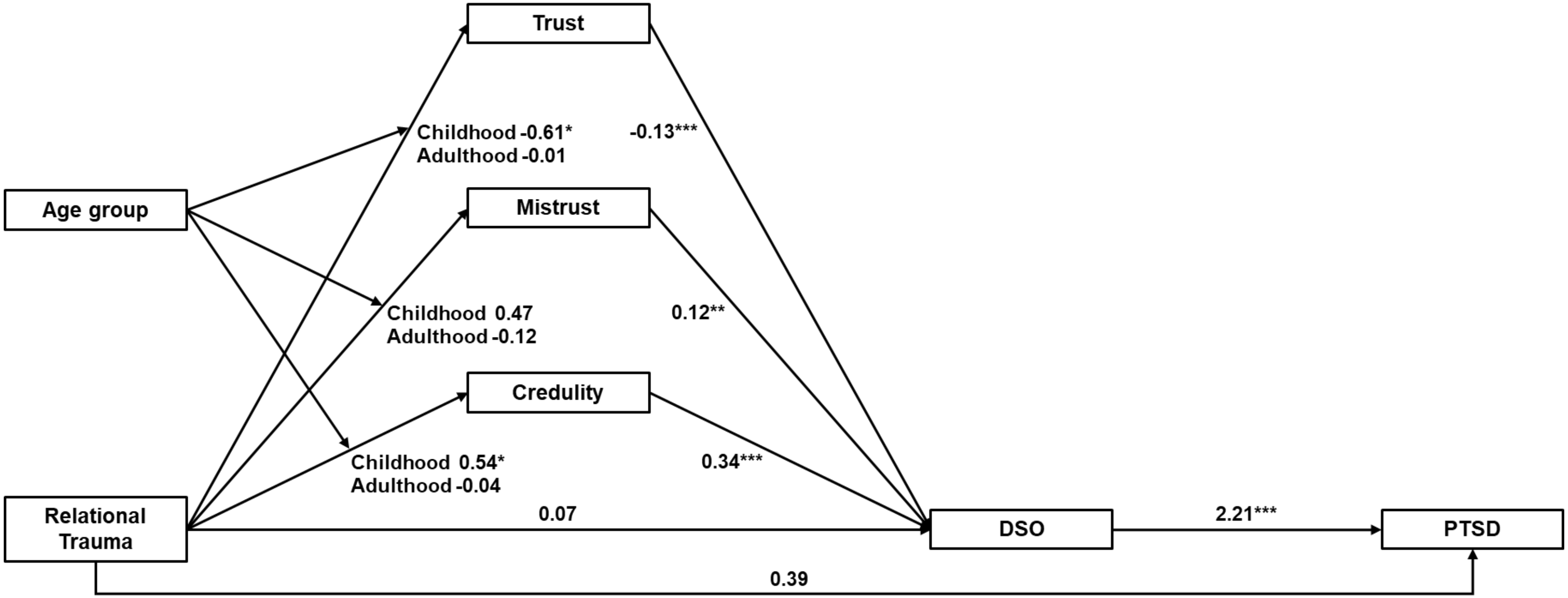

*Note.* The model was conducted with a subsample of $n = 835$ participants. Epistemic trust, mistrust, and credulity were assessed with the ETMCQ (Campbell et al., 2021). The ITQ (Cloitre et al., 2018) was used to assess symptoms of PTSD and disturbances in self-organization (DSO). Parameters were determined using a custom model in PROCESS (Hayes, 2022). Coefficients are standardized. * $p < .05$; ** $p < .01$; *** $p < .001$.
